# On the origin of Omicron’s unique Spike gene insertion

**DOI:** 10.1101/2022.06.03.22275976

**Authors:** A.J. Venkatakrishnan, Praveen Anand, Patrick J. Lenehan, Rohit Suratekar, Bharathwaj Raghunathan, Michiel J.M. Niesen, Venky Soundararajan

**Author notes:** Correspondence to: Venky Soundararajan.

## Abstract

The emergence of a heavily mutated SARS-CoV-2 variant (Omicron; B.1.1.529/BA.1/BA.2) and its rapid spread globally created public health alarms. Characterizing the mutational profile of Omicron is necessary to interpret its shared or distinctive clinical phenotypes with other SARS-CoV-2 variants. We compared the mutations of Omicron with prior variants of concern (Alpha, Beta, Gamma, Delta), variants of interest (Lambda, Mu, Eta, Iota and Kappa), and ∼1500 SARS-CoV-2 lineages constituting ∼5.8 million SARS-CoV-2 genomes. Omicron’s Spike protein has 26 amino acid mutations (23 substitutions, two deletions and one insertion) that are distinct compared to other variants of concern. Whereas the substitution and deletion mutations have appeared in previous SARS-CoV-2 lineages, the insertion mutation (ins214EPE) has not been previously observed in any other SARS-CoV-2 lineage. Here, we discuss various mechanisms through which the nucleotide sequence encoding for ins214EPE could have been acquired and highlight the plausibility of template switching via either the human transcriptome or prior viral genomes. Analysis of homology of the inserted nucleotide sequence and flanking regions suggests that this template switching event could have involved the genomes of SARS-CoV-2 variants (e.g. B.1.1 strain), other human coronaviruses that infect the same host cells as SARS-CoV-2 (e.g. HCoV-OC43 or HCoV-229E), or a human transcript expressed in a host cell that was infected by the Omicron precursor. Whether ins214EPE impacts the epidemiological or clinical properties of Omicron (e.g. transmissibility) warrants further investigation. There is also a need to understand whether human host cells are being exploited by SARS-CoV-2 as an ‘evolutionary sandbox’ for inter-viral or host-virus genomic interplay to produce new SARS-CoV-2 variants.

## Introduction

A new SARS-CoV-2 variant with an extensively mutated Spike protein was first reported to the World Health Organization (WHO) from South Africa on November 24, 2021, with the first sample collected on November 9, 2021. This strain has since been denoted as the Omicron variant (WHO nomenclature) and B.1.1.529/BA.1/BA.2 (PANGO lineage)^1^. The rapid assessment of the variant by *The Technical Advisory Group on SARS-CoV-2 Virus Evolution* and classification of Omicron as a variant of concern by the WHO within 48 hours facilitated timely epidemiological surveillance. After its initial discovery, this variant rapidly spread across the globe and was detected in over 75 countries across six continents by December 16, 2021.^2,3^

Thoroughly characterizing the mutational profile of Omicron is the necessary first step to begin interpreting its shared or distinctive clinical phenotypes, sensitivity or resistance to existing vaccines, and whether Omicron-like variants that evolve in the future may have heightened virulence. Indeed, SARS-CoV-2 has evolved into different variants of concern and variants of interest through a combination of missense, deletion, insertion, and other mutations. For example, the D614G substitution in the Spike (S) protein, which emerged early and has been detected in nearly all SARS-CoV-2 genomes in GISAID since mid 2020, increases the replication capacity and infectivity of SARS-CoV-2.^4,5^ Other substitutions (e.g. E484K and E484A) have led to significant changes in the Spike-ACE2 binding affinity, and deletions (e.g. ΔY144) have modulated the effect of neutralizing anti-Spike antibodies.^6–13^ Insertion mutations have been less prevalent in SARS-CoV-2 evolution.^14^ However, one of the most functionally consequential mutations in the evolutionary history of SARS-CoV-2 till date was the “PRRA” Spike protein insertion in the S1/S2 cleavage site, which introduced the polybasic FURIN cleavage site that mimics the RRARSVAS peptide in human ENaC-alpha.^15–19^ This insertion serves as an important determination of SARS-CoV-2 transmission, at least in part by facilitating an endosome-independent entry pathway into respiratory epithelial cells which bypasses important innate antiviral responses.^18^ It is also mechanistically required for the syncytium-mediated lymphocytes death which may contribute to the lymphopenia that is often clinically observed in COVID-19 patients.^20–23^ This insertion is an important determinant for SARS-CoV-2 transmission The availability of 5.8 million SARS-CoV-2 genomes covering ∼1500 lineages from over 203 countries/territories in the GISAID database from the beginning of the pandemic gives an opportunity to characterize the mutational profile of the Omicron variant in comparison to other SARS-CoV-2 variants.

In this study, we compare the mutational profile of Omicron with all other SARS-CoV-2 lineages including the variants of concern and variants of interest. We highlight that Omicron’s Spike protein harbors an insertion mutation ins214EPE that is absent in all other SARS-CoV-2 lineages. Given the salience of viral genetic recombination and the debated plausibility of host genome integration by SARS-CoV-2^24–26^, we considered a variety of host-viral and inter-viral genomic matter exchange scenarios that may have contributed to the adoption of this insertion mutation in the precursor variant of Omicron. We discuss potential sources for the origin of the ins214EPE and highlight the need to experimentally characterize the role of ins214EPE for transmission and immune evasion.

## Results

### Comparison of mutations in Omicron to previous SARS-CoV-2 lineages shows the presence of a unique insertion mutation in Omicron’s Spike protein

The Omicron variant harbors 37 mutations in the Spike protein, which include six deletion mutations, one insertion mutation and 30 substitution mutations.^27^ 16 of the 37 mutations are surge-associated mutations^28^ (**Table 1;** see **Methods**), i.e. their mutational prevalence increased during any three-month window when COVID-19 cases surged. Comparing these Spike protein mutations in Omicron with pre-existing variants of concern (VOCs: Alpha, Beta, Gamma and Delta) shows that 26 mutations are distinct to Omicron and seven mutations overlap between Omicron and Alpha (**Figure 1A**). When compared to the Delta variant, the mutational load of Omicron is particularly high in the Spike protein sequence, with more similar rates of mutation in regions of the viral genome encoding other proteins (**Figure S1**).

**Table 1.**
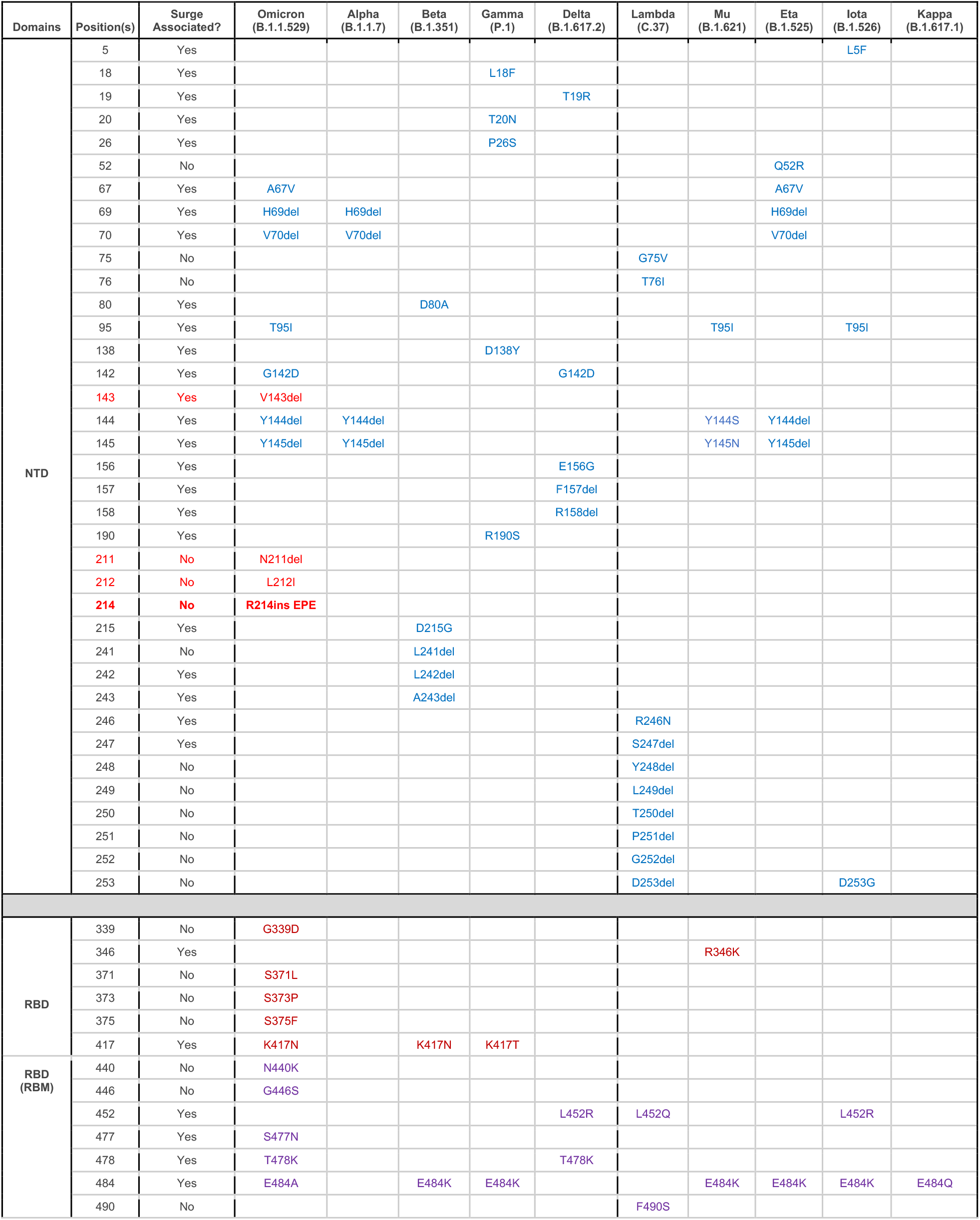
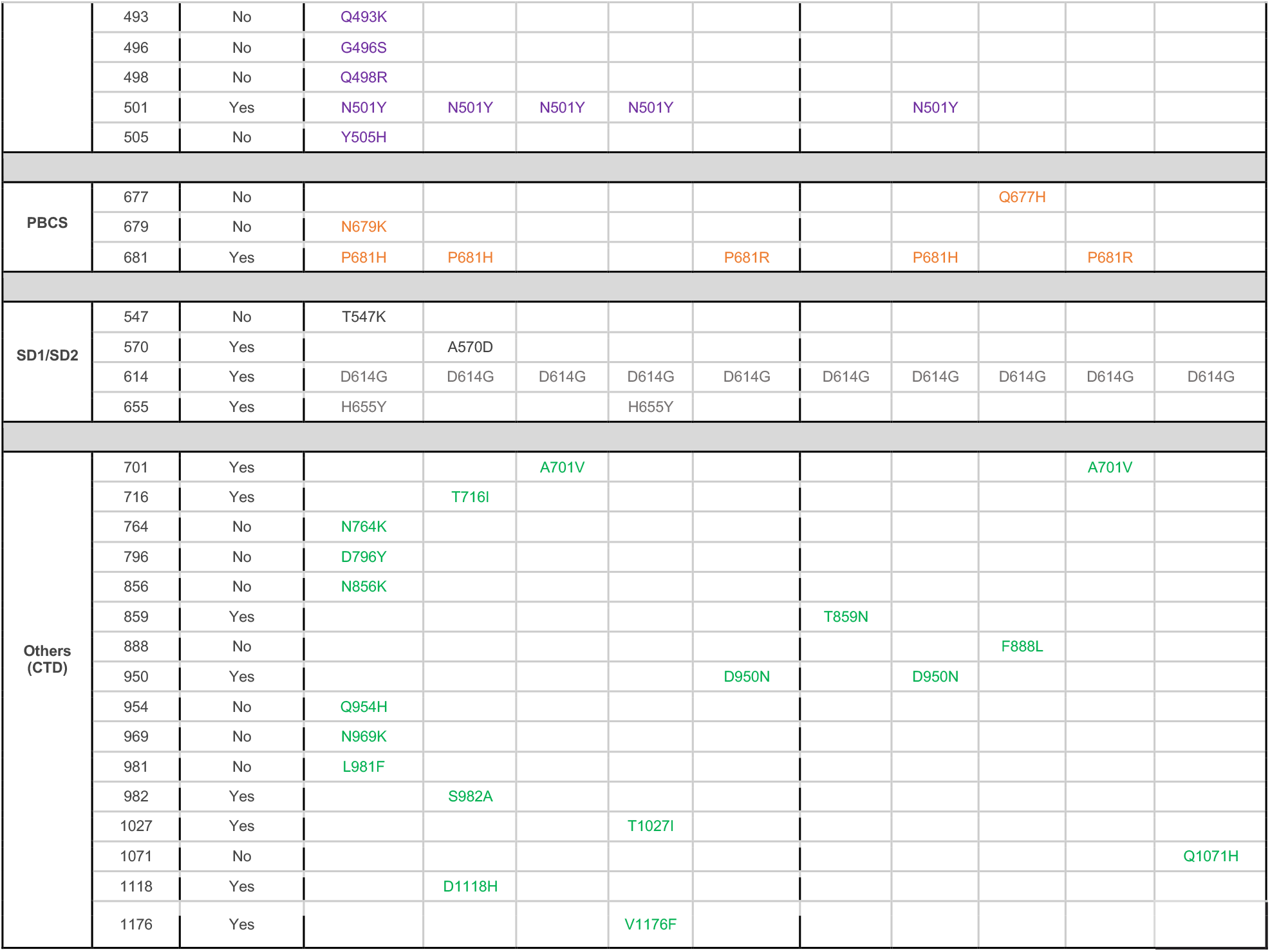
Comparison of the mutations between the Omicron variant and previously identified variants of interest/concern. The first column denotes the protein domain where the mutation is present. NTD represents the N-terminal domain; RBD represents the receptor binding domain (RBM represents the receptor binding motif); PBCS denotes poly-basic cleavage site and CTD denotes C-terminal domain.

**Figure 1.**
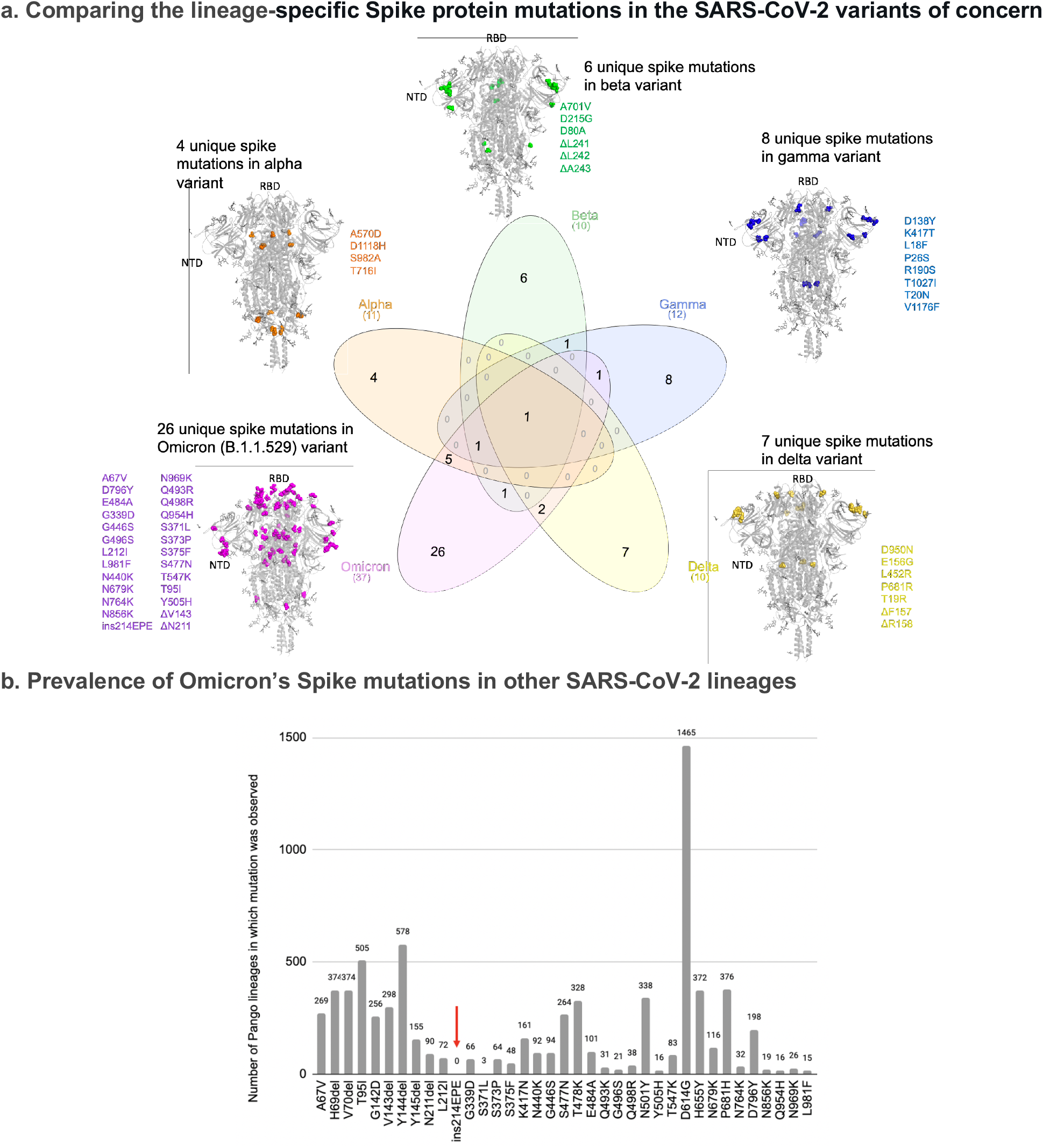
Venn diagram depicting the overlap of lineage specific spike mutations in the SARS-CoV-2 variants of concern. (A) The unique mutations observed in the spike protein for each of the variants are highlighted (spheres) on the homo-trimeric Spike protein of SARS-CoV-2. The Omicron (B.1.1.529/BA.1/BA.2) variant has the highest number (26) of unique mutations in the spike protein from this perspective, making its emergence a “step function” in evolution of SARS-CoV-2 strains. (B) Barplot denoting the number of SARS-CoV-2 lineages (besides Omicron) where the mutations present in Omicron are observed. Red arrow highlights the insEPE214 mutation which is absent from all other SARS-CoV-2 lineages.

We next analyzed which of the 26 Omicron mutations appeared in the prior variants of interest (VOIs: Lambda, Mu, Eta, Iota and Kappa) or prior SARS-CoV-2 lineages by comparing with mutations from 5,781,715 genomes corresponding to ∼1500 lineages from the GISAID database (GISAID) (**Table 1; Table S1**). Interestingly, while other insertions at or near this position have been observed in other lineages,^14,29^ the specific insertion mutation ins214EPE (**Figure S2**) has not been previously observed in any SARS-CoV-2 lineage other than Omicron (**Figure 1B**). Specifically, of the 1168 SARS-CoV-2 genomes in GISAID harboring this insertion, 1164 are classified as Omicron. Of the remaining four genomes, three have not yet been assigned a Pango lineage, while the other one (which was deposited on November 24, 2021) is labeled as B.1 (a parent lineage of Omicron). On the other hand, all of the Omicron substitution and deletion mutation have appeared in previous SARS-CoV-2 lineages (**Figure 1B, Table S1**).

The EPE insertion (Ins214EPE) on Omicron maps to the Spike protein N-terminal domain (NTD) distal from the antibody binding supersite.^11^ However, the loop where the insertion is present maps to a known human T-cell epitope on SARS-CoV-2.^30^ Further studies will be necessary to understand whether this insertion may help SARS-CoV-2 escape T-cell immunity.^14^ A recent study also suggests that insertions in the NTD, including ins214EPE in Omicron, may increase viral transmissibility by enhancing sialic-acid receptor binding.^31^ Such a functional consequence may be consistent with the observation that multiple SARS-CoV-2 lineages have acquired insertions of 1 to 8 additional amino acids at or near this same site (i.e. between positions 213 and 216).^29^ Given the well-established potential for insertions to impact SARS-CoV-2 virulence (e.g. the PRRA insertion giving rise to a polybasic FURIN cleavage site in the original SARS-CoV-2 strain), it is important to understand the functional significance and evolutionary origins of the ins214EPE insertion in the Omicron variant.^15–17,19^

### Template switching is a plausible mechanism for the origin of ins214EPE in Omicron

Ins214EPE is an in-frame insertion of 9 nucleotides that occurred between nucleotide positions 22204 and 22207. Based on the local sequence alignment, there are three candidate insertions: (1) 5’-GAGCCAGAA-3’ between 22204 and 22205, (2) 5’-AGCCAGAAG-3’ between 22205 and 22206, and (3) 5’-AGCCAGAAG-3’ between positions 22206 and 22207 (**Figure S2**). According to a recent analysis of the secondary structure of the reference SARS-CoV-2 genome, the nucleotides between positions 22205 and 22208 in (5’-GAUC-3’) constitute an RNA loop^32^, which is notable given that RNA loops appear to be more prone to insertions than stems.

A previous analysis of sequences deposited in GISAID concluded that insertions in the SARS-CoV-2 genome most likely arise from one of three mechanisms: (1) local duplications, (2) polymerase slippage, or (3) template switching.^14^ While local duplications were found to explain some short (<9 nucleotides) and long (>= 9 nucleotides) insertions, the observation that these groups had distinct nucleotide compositions, phyletic patterns, and genomic localizations led to the conclusion that short and long insertions typically arise from distinct mechanisms.^14^ Specifically, it was suggested that polymerase slippage is the best explanation for most short insertions, which typically have an excess of uracil nucleotides and are non-monophyletic. It is proposed that slippage is most likely to occur during runs of uracils owing to the slow processing of polyU tracts that has been demonstrated for the RNA-dependent RNA polymerase (RdRp) of SARS-CoV-1 and is hypothesized to be true for the RdRp of SARS-CoV-2 as well.^14,33^ On the other hand, it was posited that long insertions typically arise from template switching, given that their nucleotide composition is consistent with the SARS-CoV-2 genome, they are typically monophyletic, and they tend to occur at or near sites that were previously identified as hotspots for template switching.^14,34^ It is notable that template switching is a normal part of the life cycle for *Coronaviridae*, as discontinuous transcription via template switching is responsible for the synthesis of subgenomic RNAs (sgRNAs).^35,36^ In this light, we asked which mechanism is the most fitting explanation for the origin of ins214EPE in Omicron.

As mentioned above, exact duplication of an adjacent nucleotide sequence has been observed previously as a mechanism for both short and long insertions in the SARS-CoV-2 genome.^14^ For example, in three GISAID sequences of the B.1.429 lineage, there is a duplication of 24 nucleotides (5’-AAAAGAAGAAGGCTGATGAAACTC-3’) resulting in the sequence 5’-*AAAAGAAGAAGGCTGATGAAACTC***AAAAGAAGAAGGCTGATGAAACTC**-3’ at position 29387 (corresponding to the Nucleocapsid protein amino acid position 372).^14^ However, the inserted nucleotide sequence resulting in the Omicron ins214EPE is not a result of such a local duplication, as it is not identical or closely homologous to the preceding or subsequent nucleotide sequences in the original reference genome sequence of SARS-CoV-2 or that of the Omicron variant (**Figure S2**).

We thus asked whether polymerase slippage or template skipping was a more plausible explanation for the origin of ins214EPE. For several reasons, it appears that template switching is the more plausible hypothesis. First, this is a long insertion per the definition described previously (≥9 nucleotides), and long insertions are more likely to arise from template switching than polymerase slippage.^14^ That said, we recognize that with exactly 9 nucleotides, this is a borderline case between short and long insertions, and so its length alone may have limited value in distinguishing between these mechanisms. Second, this insertion has no uracil nucleotides, contrary to the expected excess uracils in slippage-mediated insertions.^14^ Third, this insertion is monophyletic, although it is worth noting that other insertions at the same location have been observed in several other SARS-CoV-2 lineages.^14,29^ Finally, the insertion occurs near previously described sites of potential non-canonical template switching. Specifically, there were non-canonical junctions observed 30 nucleotides upstream and 60 nucleotides downstream of this site (at positions 22183 and 22276, respectively).^34^

### Candidate templates for the origin of ins214EPE in Omicron

It is of interest to determine candidate RNA molecules from which SARS-CoV-2 template switching-mediated insertions have arisen. We reasoned that there are broadly three categories of most likely templates: (1) genomic material of SARS-CoV-2 itself (i.e., the positive-sense genomic RNA or the negative-sense anti-genomic RNA), (2) genomic or anti-genomic material of other viruses that have the capacity to co-infect the same cells as SARS-CoV-2; and (3) human transcripts that are expressed in cells infected with SARS-CoV-2 (**Figure 2A-B**). Although the last category is not a well-described method of template switching,^37^ it has been suggested previously that insertions in SARS-CoV-2 genomes could be derived from the host transcriptome.^38^ We identified exact matches for the forward and/or reverse complement sequences in all three categories (**Table 2, Figure 2C**).

**Table 2.** Number of viral genomes and human transcripts with forward or reverse complement matches to the three potential insertion sequences. Because the insertion sequences occur by definition in Omicron genomes, counts are shown separately for total SARS-CoV-2 genomes from GISAID and genomes which are not assigned to the Omicron lineage. We confirmed that no sequences that were assigned to the Omicron lineage contained the insertion sequence or its reverse complement at any sites other than at the insertion itself. For the Human *Coronaviridae* genomes, counts were first obtained by considering the available sequences for all human-infecting coronaviruses and then filtered to retain only the viruses that are known to cause common colds or enteric illness (i.e. Severe Acute Respiratory Syndrome and Middle East Respiratory Syndrome virus sequences were excluded).

| Candidate Insertion Sequence | Human Transcriptome |  | SARS-CoV-2 Genomes from GISAID |  | Human *Coronaviridae* Genomes |  |
| --- | --- | --- | --- | --- | --- | --- |
|  | Transcripts (Genes) |  | Total (Non-Omicron) |  | Any (Seasonal or Enteric) |  |
|  | Forward | Rev. Comp | Forward | Rev. Comp | Forward | Rev. Comp |
| 5'-GAGCCAGAA-3' | 4677<br>(1534) | 3264<br>(1220) | 2100<br>(931) | 27<br>(27) | 18<br>(8) | 17<br>(6) |
| 5'-AGCCAGAAG-3' | 6190<br>(2008) | 4293<br>(1591) | 1275<br>(106) | 269<br>(269) | 3<br>(0) | 12<br>(0) |
| 5'-GCCAGAAGA-3' | 5210<br>(1564) | 3146<br>(1144) | 1319<br>(150) | 201632<br>(201632) | 13<br>(7) | 4<br>(2) |

**Figure 2.**
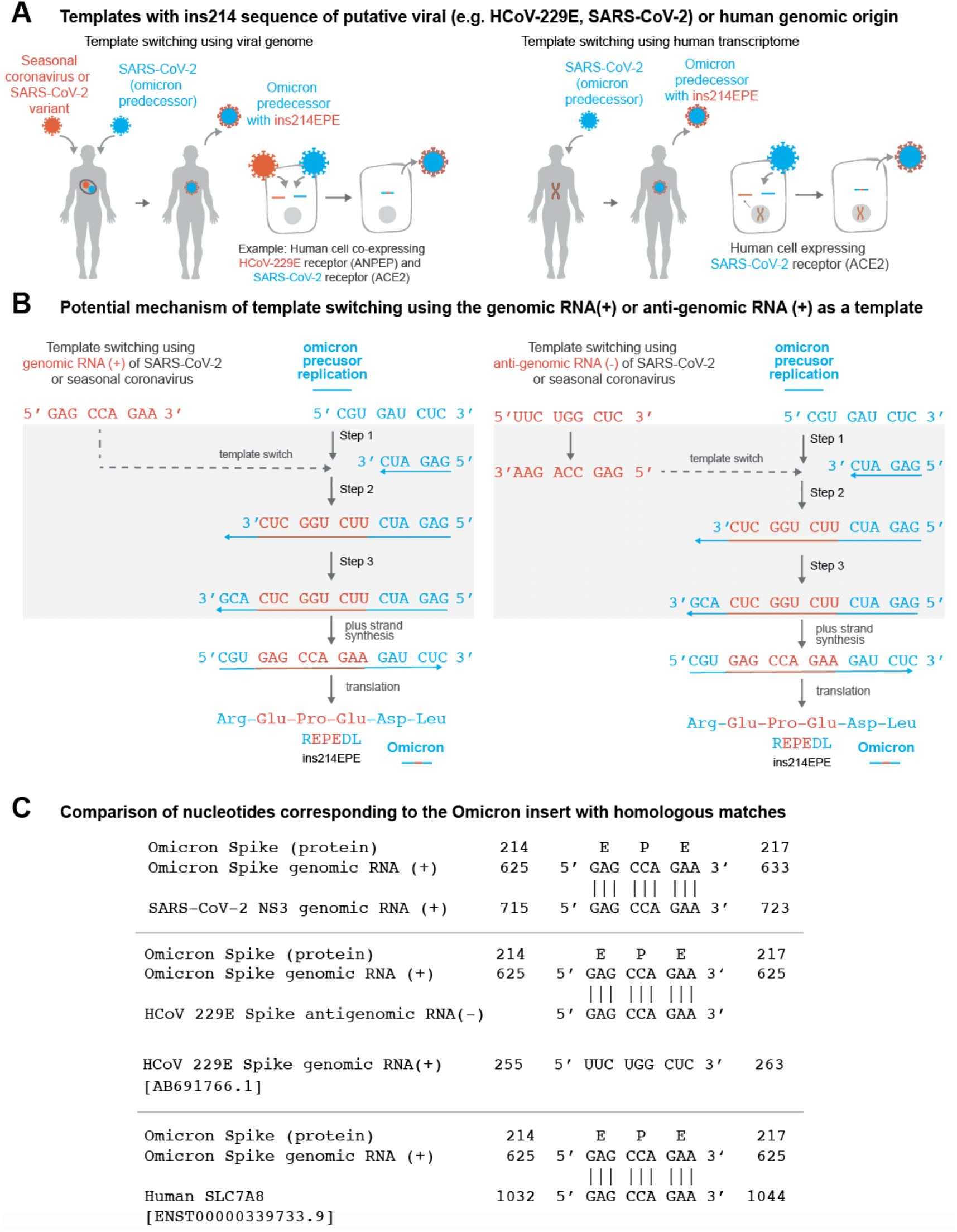
(A) Schematic representation of Omicron evolution through template switching involving viral (e.g. seasonal coronavirus or SARS-CoV-2) or human RNA. (B) Potential mechanism of template switching using viral genomic RNA (positive sense) or anti-genomic RNA (negative sense) as a template. Step 1: Negative strand synthesis begins using Omicron predecessor’s genomic RNA as template. Step 2: Negative strand synthesis temporarily uses the genomic or anti-genomic RNA of SARS-CoV-2 or a co-infecting virus. Step 3: Negative strand synthesis resumes using Omicron predecessor’s genomic RNA as template. (C) Examples of matches identical to the nucleotide sequence ‘GAG CCA GAA’ in the SARS-CoV-2 genome, the HCoV-229E anti-genome, and a human SLC7A8 transcript are shown.

There are no exact matches in any Omicron genomes collected to date for the three candidate sequences outside of the insertion site itself. This is notable because in previous template switch-mediated insertions in the SARS-CoV-2 genome, the putative insertion template (“origin”) has typically been present in the insertion-containing genome (**Table S2**).^14^ For the Omicron insertion, on the other hand, the only SARS-CoV-2 sequences in GISAID containing exact forward or reverse complement matches are assigned to other SARS-CoV-2 lineages (or were not assigned to any lineage at the time of this analysis). There are several possible implications or interpretations of this finding.

First, substitutions can be introduced within the inserted sequence during template switching itself or during subsequent rounds of viral replication, which would result in imperfect matching between the insertion and template sequences. This is particularly relevant for Omicron, which represents a long phylogenetic branch that arose after presumably several months of unobserved evolution.^39,40^ We indeed found that the reference Omicron genome (EPI_ISL_6640916)^39^ harbors several nucleotide 9-mers that differ from the candidate insertions by only a single nucleotide (**Table S3**). These should be considered as possible templates for ins214EPE. Second, it is possible that the utilized template was derived from a co-infecting SARS-CoV-2 variant that does harbor the exact inserted sequence. Recombination between SARS-CoV-2 lineages in the context of simultaneous co-infection has been described previously, with particularly high recombination rates seen in the Spike protein sequence.^35,41^ The distributions of lineages comprising the matched genomes for each candidate insertion are shown in **Table S4**. Finally, we noticed that several of the non-Omicron genomes with exact matches to one or more of the candidate insertions have been assigned to the B.1.1 Pango lineage (or sublineages thereof) (**Table S4**). Given that Omicron (originally Pango lineage B.1.1.529) is a phylogenetic descendant of B.1.1, this suggests that the ancestral genome which evolved into Omicron could have provided the necessary template for this insertion.

We also identified several genomes (or anti-genomes) of seasonal or enteric human-infecting coronaviruses that contain one or more of the putative insertion sequences. For example, multiple human coronavirus OC43 (HCoV-OC43) and human enteric coronavirus strain 4408 genomes both contain 5’-GCCAGAAGA-3’ and 5’-GAGCCAGAA-3’ in their nucleocapsid and replicase polyprotein genes, respectively (**Table S5**). Further, 5’-GAGCCAGAA-3’ was present in the 33 HCoV-229E anti-genomes, and 5’-GCCAGAAGA-3’ was present in two HCoV-NL63 anti-genomes (**Table S5**). The importance of recombination between coronaviruses has been highlighted recently,^42^ and its potential is supported by clinical reports showing that COVID-19 patients are co-infected with other respiratory pathogens, including non-SARS-CoV-2 viruses of the *Coronaviridae* family, at relatively high frequencies.^43–45^ Further, host receptors utilized by other coronaviruses (e.g. ANPEP and DPP4) are co-expressed with the SARS-CoV-2 receptor (ACE2) at the single cell level in respiratory and/or gastrointestinal epithelial cells (**Tables S6–S7**),^46^ which could facilitate co-infection at the cellular level (a prerequisite for genomic recombination). Intestinal co-expression is relevant given the evidence that SARS-CoV-2 and other coronaviruses can infect enterocytes.^47–49^ That said, it is worth noting the possibility that genomic material of pathogens outside of the *Coronaviridae* family (perhaps even non-viral pathogens) could also serve as substrates for template switching in the context of such cellular co-infection.

Finally, there were 4677 human transcripts (from 1534 genes) containing the forward sequence 5’-GAGCCAGAA-3’ and 3264 human transcripts (from 1220 genes) containing the reverse complement sequence. Similar summary statistics are shown for the two other potentially inserted 9-nucleotide sequences in **Table 2**.

### Consideration of local homology for the candidate templates

This landscape of possible templates, particularly among human transcripts, is expectedly quite vast given the reasonably small total space of possible 9-mer nucleotide combinations (4^9^ = 262144). This raises the question of whether the most likely candidates may be those transcripts which have more homology surrounding the inserted sequence, as complementary base pairing resulting from such local similarity can increase the likelihood of serving as a template for recombination.^37^ Indeed, in the normal process of coronavirus genomic replication, the prevailing model is that subgenomic RNAs are generated during negative strand synthesis via a template switching mechanism that relies on homology between conserved transcription regulatory sequence (TRS) elements dispersed strategically throughout the genome.^34,36,50,51^ However, in the context of SARS-CoV-2, non-canonical transcripts with junctions that are not derived from TRS sequences and that share little homology between the 5’ and 3’ sites suggests that template switching guided by partial complementarity or other mechanisms may also play a role.^34,52^

To study the homology between the regions surrounding template switch-mediated insertions and their origins (i.e. the putative template that was copied to generate the insertion), we first considered a “positive control” set of four 12-15 nucleotide insertions in SARS-CoV-2 that were previously attributed to template switching with high confidence (**Table S2**).^14^ We calculated a normalized homology score (NHS) between the 7 or 35 nucleotides upstream or downstream of the insertion and origin sequences in these genomes (see **Methods**). Surprisingly, the degree of homology observed was generally not higher than expected by chance, as assessed via the NHS distribution of 10,000 randomly paired non-overlapping 7-mer or 35-mer nucleotide sequences from the SARS-CoV-2 genome (**Figure S3, Table S8**). This suggests that local homology may not be a prerequisite for the generation of template switch-mediated insertions in SARS-CoV-2.^34^

Nevertheless, we still assessed the homology between the 35 nucleotides upstream or downstream of the Omicron insertion and the 35 nucleotides upstream or downstream of all candidate templates (see **Methods**). The NHS distributions for these candidates were generally similar to the distribution observed previously for randomly selected SARS-CoV-2 35-mers (**Figure S4**). Candidates in each category with the highest degrees of homology in the flanking upstream or downstream sequences included SARS-CoV-2 genomes from the lineages B.1.609 (NHS = 69) and AY.103 (NHS = 66), the HCoV-229E Spike protein (NHS = 63), and human transcripts of ACTN1 (NHS = 74) and EMC4 (NHS = 71). Some candidate templates also showed more homology in shorter sequences directly upstream and/or downstream of the inserted sequence. For example, in the reverse complement of the human TMEM245 transcript, there is a 17-nucleotide stretch with exact homology to the insertion-containing region of the Omicron genome (i.e. the 9-nucleotide inserted sequence plus 8 exactly matched flanking nucleotides) (**Figure S5**).

## Discussion

Omicron is more highly transmissible than prior variants,^53,54^ is less susceptible to neutralization by monoclonal antibodies and sera of vaccinated individuals,^55–61^ and is more likely to cause re-infections and vaccine breakthrough infections.^62–64^ Among the many mutations in its Spike protein, ins214EPE is the only one that has not previously been observed in any other lineages. Whether this insertion, singularly or in concert with other mutations, contributes to high transmissibility or lower susceptibility of neutralization by antibodies warrants further investigation.

While it is challenging to definitively determine the mechanism that gave rise to ins214EPE, we propose that template switching is a plausible explanation (**Figure 2**). Although the RNA-dependent RNA polymerases of SARS-CoV-2 and other coronaviruses do normally utilize template switching to generate subgenomic RNAs,^34,50^ it appears that template switch-mediated insertions may derive from a non-canonical form of this process in which a high degree of local homology (e.g. homologous TRS core sequences in the leader and body regions of the genome) is not essential.^34,52^ Here we have highlighted several possible sources of the template for this insertion, including the SARS-CoV-2 genome itself along with the genomes of other viruses or human transcripts. The use of SARS-CoV-2 genomic material as the template is supported by the finding that SARS-CoV-2 genome replication occurs in organelles which spatially concentrate the viral genomic material and replication machinery.^65^ That said, it might indeed be possible for non-SARS-CoV-2 viral genomes or host mRNAs to be aberrantly included in these organelles, thus rendering them accessible for utilization during template switching as well.

There may also be other mechanisms that could lead to SARS-CoV-2 insertions in addition to those which are considered here. For example, it has been suggested that Omicron could have evolved in a non-human species such as mice or deer, in which case other viral genomes and transcripts should be considered as possible templates.^66,67^ Also, a recent analysis suggests that the proofreading exoribonuclease (encoded in nonstructural protein 14, or nsp14) is required for at least some of the genetic recombination observed in SARS-CoV-2,^68^ but the potential mechanisms described here do not account for this. Finally, it is reasonable to question whether there is a relationship between ins214EPE and the 3-nucleotide deletion (ΔN211) that occurs shortly upstream of it in most Omicron sequences. However, because most sequences in GISAID with other insertions at position 214 do not possess such neighbording deletions (**Figure S6**), we believe that this proximal deletion was not a mechanistic prerequisite for the generation of the Omicron insertion.

It is not clear whether any one of these insertion-generating mechanisms would have more far-reaching consequences than the others. Template switch offers the intriguing possibility of new SARS-CoV-2 lineages borrowing protein domains or sub-domains from previous variants, other viruses, or human proteins. However, it is worth noting that several of the possible templates that we identified for ins214EPE were derived from viral anti-genomes (i.e. from HCoV-229E or SARS-CoV-2 anti-genomes) or the reverse complement sequences of human transcripts (e.g. TMEM245). In such cases, the polypeptide encoded in the Omicron genome would differ from that encoded by the positive-sense strand of the template. The genomic location and/or amino acid content of the inserted sequence may be more important than its origin, and we thus highlight the need to characterize the functional impact of ins214EPE on the clinical and epidemiologic properties of the Omicron variant. Importantly, the data and hypotheses presented here are not sufficient to make inferences about such properties including transmissibility, immune evasion, or disease severity. Even if Omicron did acquire an insertion by utilizing the genome of a common cold-causing coronavirus (e.g. HCoV-OC43, HCoV-229E) or a host transcript, we do not propose that this would explain the reduced severity of COVID-19 observed in patients infected with Omicron compared to prior VOCs.^69–72^

The rapid rise in COVID-19 cases attributed to the Omicron variant, including among fully vaccinated individuals, raised alarm globally. In this context, it is important to better understand both the origins and consequences of new genomic alterations that distinguish Omicron from prior VOCs and VOIs. Here, we begin to address the former by providing several plausible hypotheses on the origins of a 9-nucleotide insertion in the N-terminal domain of the Omicron Spike protein. We suggest that genomic surveillance strategies should include an emphasis on sequencing SARS-CoV-2 genomes from immunocompromised patients and individuals with viral co-infections (including co-infections with multiple SARS-CoV-2 variants), as such individuals may provide unique contexts for genomic recombination and the evolution of new variants.

## Methods

### Analysis of surge-associated mutations

Core mutations were derived from parental lineages from the CoV-RDB database for each Variant of Interest or Variant of Concern: Alpha (B.1.1.7), Beta (B.1.351), Gamma (P.1), Delta (B.1.617.2), Lambda (C.37), Mu (B.1.621), Eta (B.1.525), Iota (B.1.526), Kappa (B.1.617.1) and Omicron (BA.1).^27^ All SARS-CoV-2 genome sequences corresponding to the Omicron variant were directly derived from the GISAID database.^2,3^ There were 1448 SARS-CoV-2 genomes annotated as B.1.1.529/BA.1/BA.2 as of December 13, 2021).

To understand which SARS-CoV-2 mutations correlate with COVID-19 test positivity, we identified “surge-associated mutations” using genomic data from GISAID (5,781,715 sequences from 203 countries/territories between December 2019 to December 2021) and epidemiology data from Our World in Data (OWID)^73^ as described in our previous study.^28^ A mutation is considered “surge associated” if it satisfies the following criteria: (1) it is present in at least 100 SARS-CoV-2 sequences in GISAID; (2) in a period of three consecutive months during which there was a monotonic increase of PCR positivity by at least 5% in a given country, the prevalence of the mutation in that country also monotonically increased by at least 5%.

### Nucleotide 9-mer search to identify candidate viral and human templates for ins214EPE

A exact 9-mer search for all the three possible inserts (5’-AGCCAGAAG-3’, 5’-GAGCCAGAA-3’, 5’-GCCAGAAGA-3’) and their reverse complement sequences was performed across three different databases: (i) human transcriptome, (ii) SARS-CoV-2 genomes from GISAID, and (iii) human-infecting *Coronaviridae* family viruses. For a reference Omicron genome (EPI_ISL_6640916),^39^ we also performed a modified search to identify all 9-nucleotide sequences that differed from the possible insertion sequences by only one nucleotide (i.e. allowing for a single mismatch). The human transcript sequences (n = 244,939) were downloaded from the Gencode database^74^ (version 39; GRCh38.p13 of human genome). Coding sequences (CDS) for 5,781,715 SARS-CoV-2 genomes were accessed from GISAID^2^ on 13th December, 2021. All available sequences from human-infecting *Coronavirdae* family (taxid:11118) viruses were accessed from the NCBI virus database ^75^ (https://www.ncbi.nlm.nih.gov/labs/virus on 13, December 2021 (n = 574,178 complete genomes with 4,028,478 CDS). In the final analysis, we excluded genomes that were labeled as SARS-CoV, SARS-CoV-2, and Middle East Respiratory Syndrome (MERS)-CoV (or otherwise named strains of these viruses) to consider only seasonal and enteric coronavirus strains: human coronavirus (HCoV)-229E, HCoV-OC43, HCoV-NL63, and human enteric coronavirus strain 4408.

### Assessment of homology between regions flanking insertion and origin sites

For a given insertion, we defined a putative origin as the matched sequence in a viral genome, a viral anti-genome, or a human transcript. We then assessed the homology between the 35 nucleotides upstream of the insertion and the 35 nucleotides upstream of the origin, and similarly we assessed the homology between the 35 nucleotides downstream of the insertion and the 35 nucleotides downstream of the origin. To assess the similarity between any given pair of nucleotide sequences, we defined a function using Bio.pairwise2 module from biopython v1.76 that performs a global alignment of nucleotide sequence using a custom scoring scheme (i.e. +5 for a match, −4 for a mismatch, 0 for gap start and extension). This score ranges from 0 (no matches) to 175 (perfect match for all 35 nucleotides). We further defined a Normalized Homology Score (NHS), ranging from 0 to 100, which is simply calculated by dividing the Homology Score by 175 and multiplying by 100. We also applied a similar protocol to assess the homology between shorter upstream and downstream sequences (7 nucleotides), where the NHS was calculated by dividing the Homology Score by 35 and multiplying by 100.

A set of “positive control” template switch-mediated insertions was obtained from Supplementary Data 4 from the prior analysis by Garushyants et al.^14^ We filtered this table to only those insertions that were assigned a mechanism of “Template switch” and which were 12 or more nucleotides long. This set of insertions is shown in **Table S3**. For each insertion, the genomic positions of the origin and insertion sequences were provided in this table, along with the Pango lineage assigned to the genome(s) harboring the insertion. We searched SARS-CoV-2 genomes in GISAID to identify those genomes that contained the provided origin and insertion sequences at approximately the provided genomic positions. The GISAID identifiers for the corresponding insertion-containing genomes are provided in **Table S3**. For each of these genomes, we then obtained the 35-nucleotide sequences upstream and downstream of the insertion and origin. Finally, we calculated the NHS for each relevant pair of sequences: (i) the 35 nucleotides upstream of the insertion versus the 35 nucleotides upstream of the origin, and (ii) the 35 nucleotides downstream of the insertion versus the 35 nucleotides downstream of the origin. These scores, and the local alignments contributing to them, are shown for each individual genome in **Table S4**.

### Single cell analysis of coronavirus receptor coexpression

Publicly available single cell RNA-sequencing datasets were obtained and processed as described previously.^46,76^ The data is hosted at https://academia.nferx.com/dv/202011/singlecell/ and includes approximately 2.8 million cells which are derived from dozens of independent studies that cover most major human tissues. We determined the total number and percentage of cells (out of these 2.8 million total cells) expressing each gene encoding a coronavirus receptor of interest (ACE2, ANPEP, DPP4), along with the number and percentage of cells co-expressing ACE2 and ANPEP or ACE2 and DPP4. We also performed similar analyses on two specific studies of interest: (i) a study of nasopharyngeal and bronchial samples from COVID-19 patients and healthy controls (approximately 33,000 cells),^77^ and (ii) a study of ileal biopsies from Crohn’s Disease patients (approximately 136,000 cells).^78^ For these studies, we also evaluated co-expression in the cell types that showed the strongest expression of these genes. Specifically, to evaluate whether the degree of co-expression for ACE2 and ANPEP or DPP4 was more than expected by chance, we calculated the Observed to Expected Ratio of co-expression, assuming that expression of each gene is distributed randomly across the analyzed cells. Specifically, this value was calculated by dividing the co-expressing percentage by the product of the individual expression percentages and multiplying by 100.

## Data Availability

All data produced in the present work are contained in the manuscript and accessible via the GISAID and NCBI resources.

## Supplementary Material

**Figure S1:**
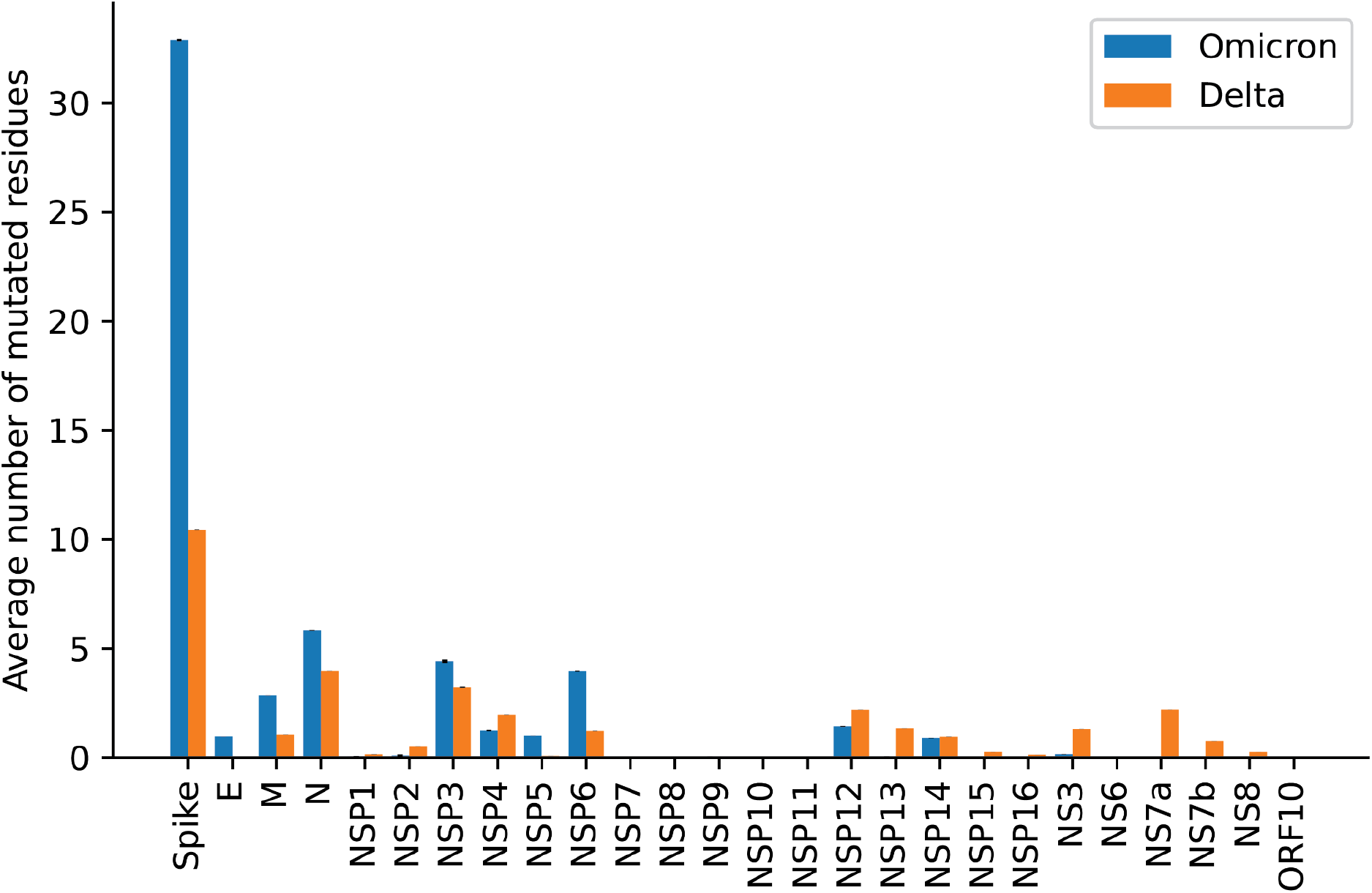
Mutational burden of all SARS-CoV-2 proteins for the Omicron variant (B.1.1.529/BA.1/BA.2) compared with the Delta variant (B.1.617.2). A total of 5,441 Omicron (B.1.1.529/BA.1/BA.2) sequences and 159,981 Delta (B.1.617.2) sequences were retrieved from the GISAID on December 16, 2021. Each bar represents the average number of mutations reported for a sequence in each of the SARS-CoV-2 proteins. Error bars represent the standard error of the mean.

**Figure S2.**
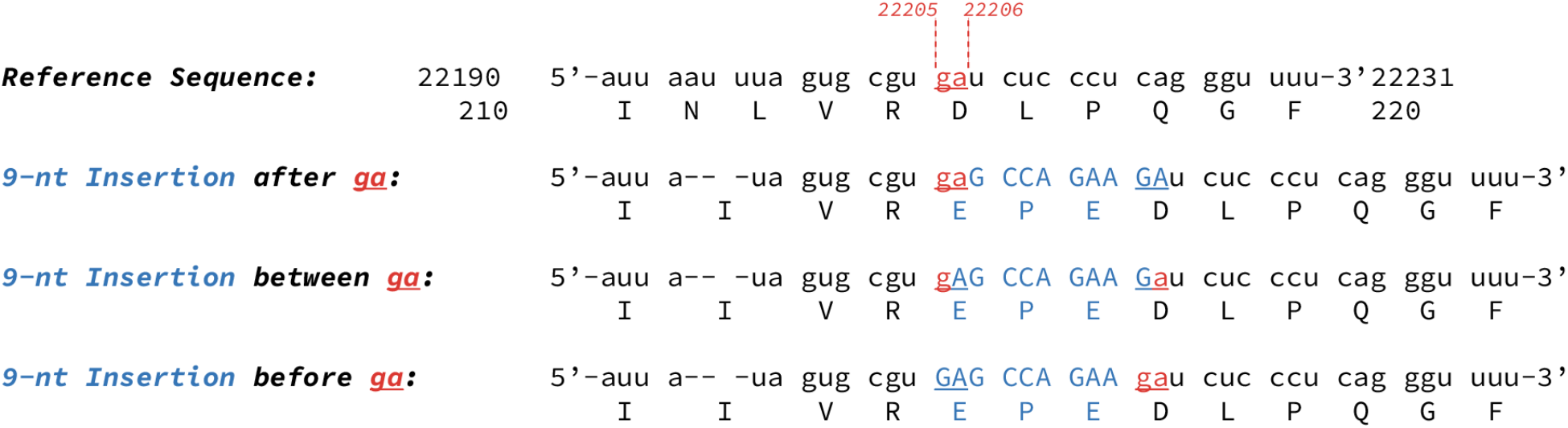
Possible nucleotide insertions giving rise to ins214EPE in the Omicron genome. There are three possible scenarios in which a 9-nucleotide insertion could yield the observed sequence. Because of the local sequence alignment (i.e. the occurrence of the insertion at a 5’-GA-3’ and the presence of a 5’-GA-3’ on either side of the insertion), it cannot be determined which scenario actually gave rise to the insertion.

**Figure S3.**
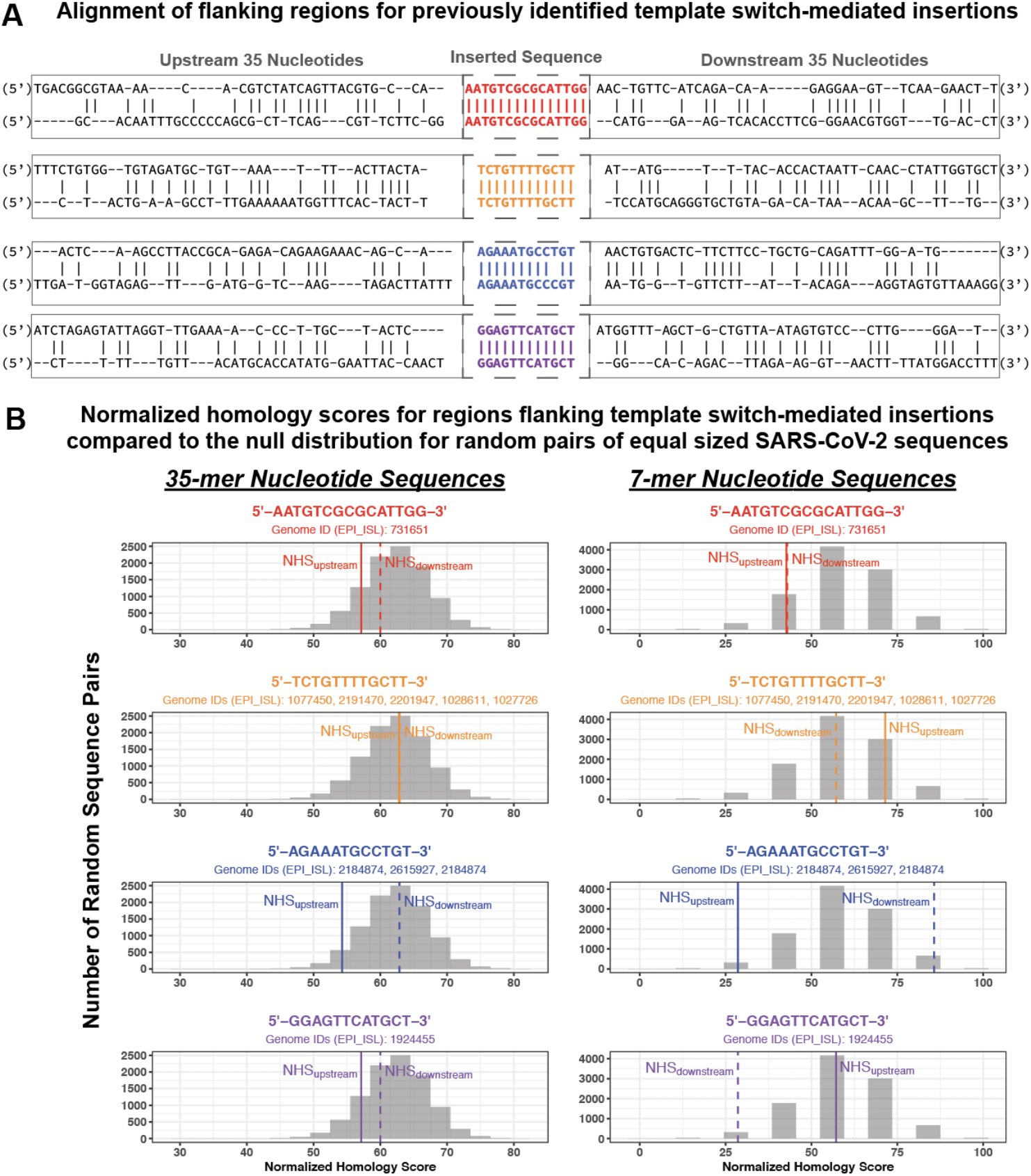
Alignment and normalized homology scores for regions flanking insertion and origin sequences for previously identified template switch-mediated insertions in the SARS-CoV-2 genome. Four “positive control” template switch-mediated insertions of 12 to 15 nucleotides were identified from the previous analysis by Garushyants et al.^14^ We identified the corresponding SARS-CoV-2 genomes in GISAID for each insertion. (A) Alignment of the insertion (top) and origin (bottom) sites for the 35 nucleotides upstream and downstream of each inserted sequence. These alignments are also shown in **Table S8** for each individual genome. (B) For a given insertion, we then calculated normalized homology scores (NHS) between the regions flanking the inserted sequence and the origin (template) sequence. Specifically, we compared the 35 nucleotides upstream of the insertion to the 35 nucleotides upstream of the origin (“NHS_upstream_”), and we separately compared the 35 nucleotides downstream of the insertion to the 35 nucleotides downstream of the origin (“NHS_downstream_”) (panel on left). We did the same to compare 7-nucleotide sequences upstream and downstream of the insertion and origin (panel on right). In each case, the normalized homology scores are shown for each insertion as vertical lines (solid line for upstream, and dashed line for downstream) on top of a control null distribution (gray histogram). The numerical scores are given in **Table S8**. To generate the null distributions of NHS values, we compared 10,000 pairs of non-overlapping 35-nucleotide (or 7-nucleotide) sequences from the original SARS-CoV-2 genome (NC_045512.2).^79^

**Figure S4.**
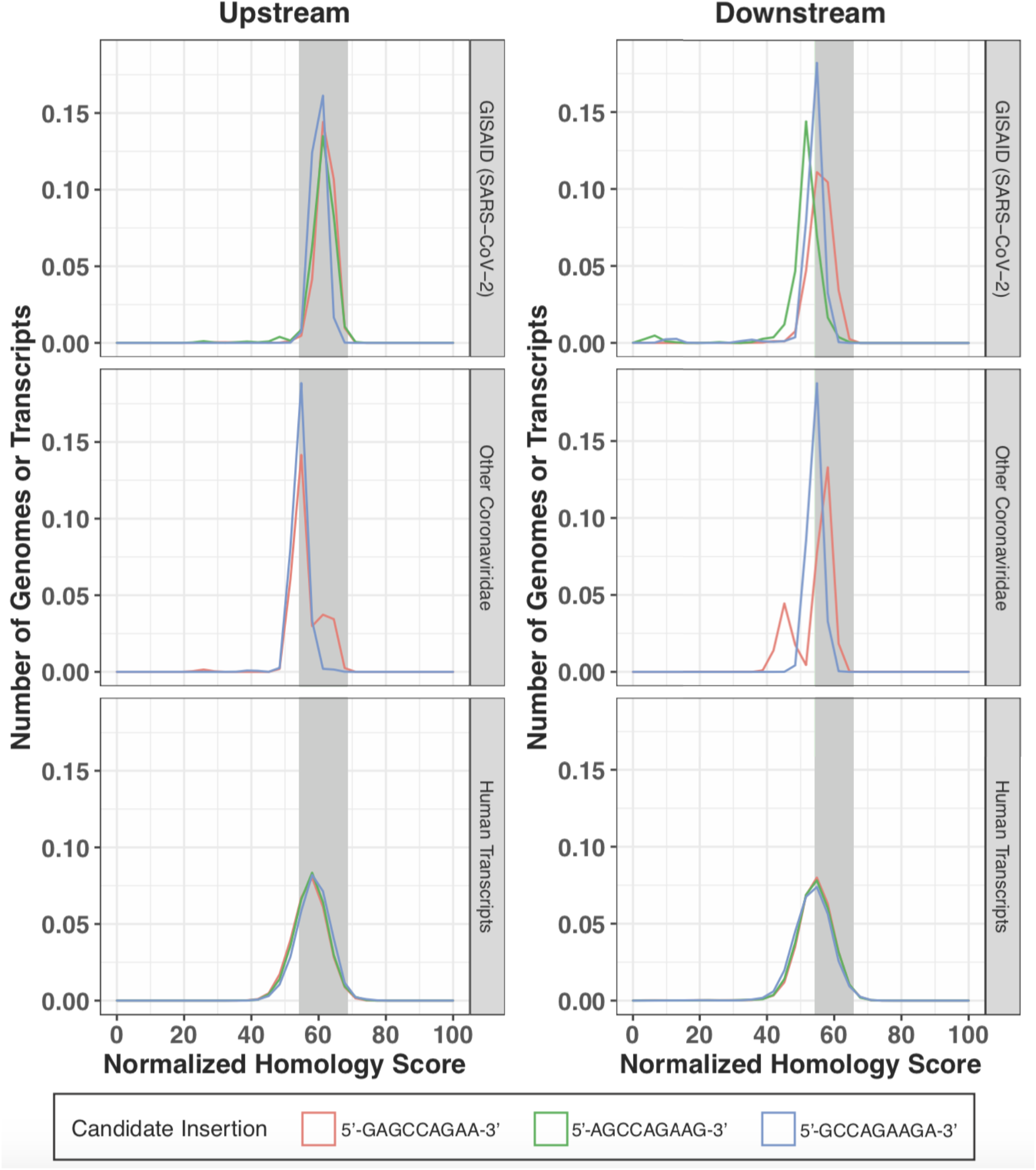
Distributions of normalized homology scores for regions flanking ins214EPE and candidate origin sequences. Distributions of normalized homology score (NHS) values between the regions flanking the Omicron insertion and the regions flanking the candidate origin sequences from SARS-CoV-2 genomes in GISAID (top), non-SARS-CoV-2 *Coronaviridae* genomes (middle), and human transcripts (bottom). Distributions are shown separately for the comparisons of nucleotide sequences upstream and downstream of the Omicron insertion and the respective putative templates. The gray box in each plot corresponds to the 5th to 95th percentile of the NHS distribution comparing randomly selected non-overlapping 35-nucleotide sequences from the SARS-CoV-2 genome (i.e. the histogram shown in each panel of Figure S3.

**Figure S5.**
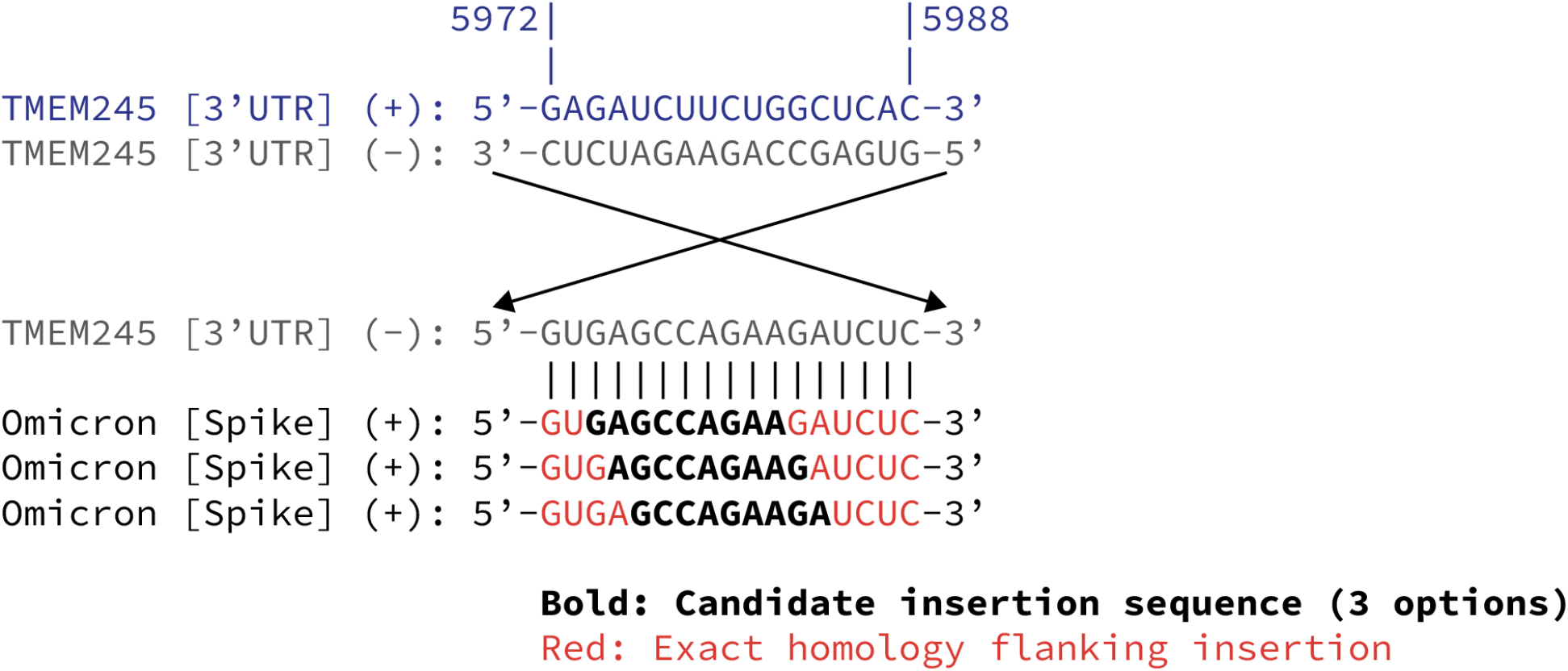
Alignment of the Omicron genomic region corresponding to ins214EPE with the human TMEM245 transcript. TMEM245 is an example of a human transcript which harbors the sequence that corresponds to ins214EPE and has exact homology in the short nucleotide sequences upstream and downstream of it. The relevant region of the TMEM245 transcript (nucleotides 5972 to 5988), which is present in the 3’ untranslated region (3’ UTR), is shown in blue. The reverse complement of this sequence is shown in gray, first in the 3’ to 5’ orientation and then flipped to visualize the 5’ to 3’ orientation from left to right. This sequence aligns exactly with the region corresponding to and surrounding ins214EPE in the Omicron genome. The three rows corresponding to the Omicron genome show identical sequences, but consider the three different candidate insertions that may have given rise to ins214EPE (see Figure S2). In each row, the candidate insertion sequence is shown in bold, and the flanking nucleotides are shown in red.

**Figure S6.**
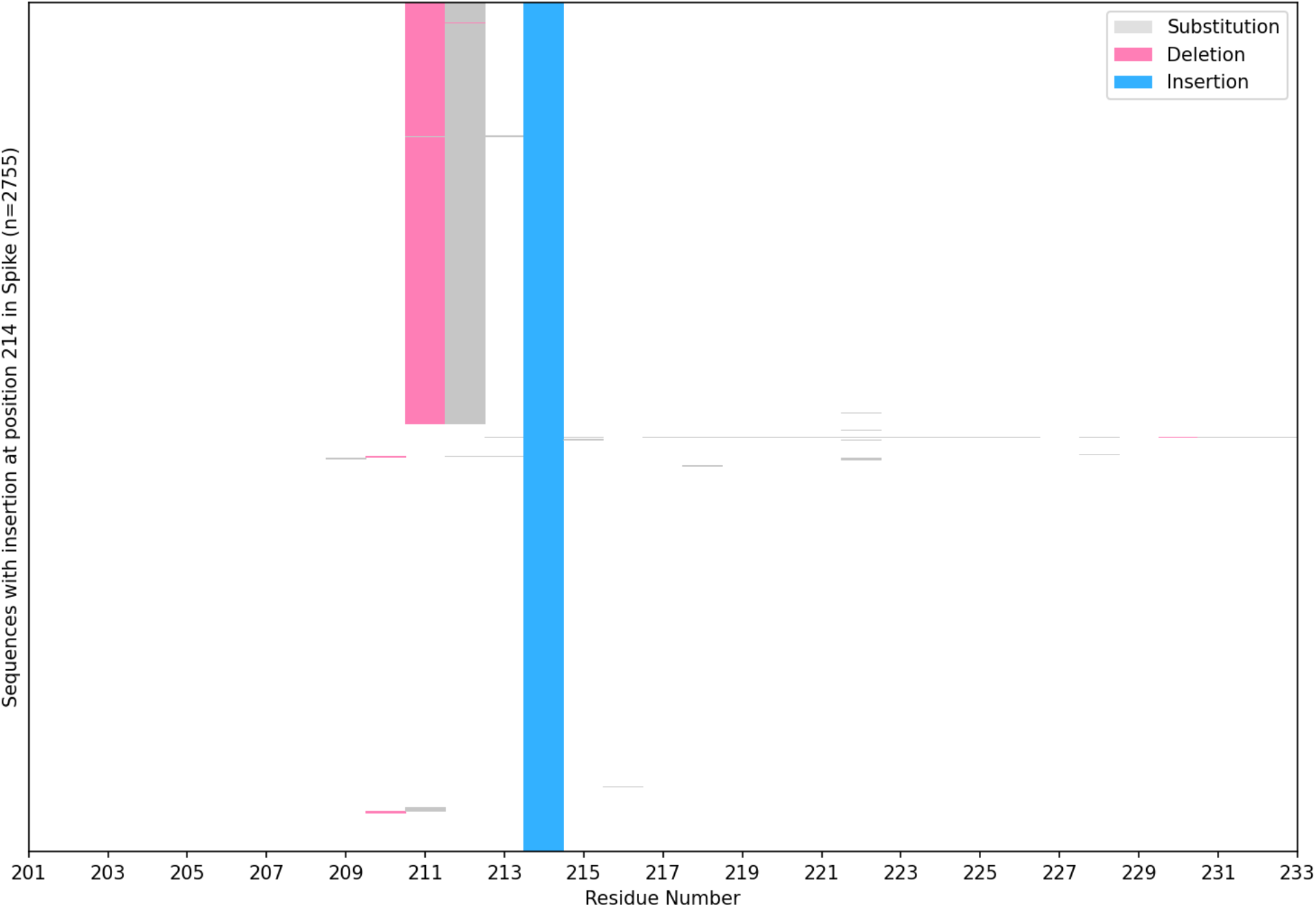
Occurrence of deletions and substitutions in the residues neighboring position 214, in SARS-CoV-2 genomes harboring any insertion at this position. Among all 2755 SARS-CoV-2 sequences in GISAID with any insertion at position 214 (as of December 14, 2021), we assessed the neighboring residues for the occurrence of deletions, substitutions, or other insertions. The sequences with a deletion observed at position 1358 all correspond to the Omicron variant (two sequences lack assigned lineages and one is assigned to a parent lineage), whereas other sequences containing alternate insertions at position 214 (i.e. not ins214EPE) generally have no alterations in the neighboring regions.

**Table S1.**
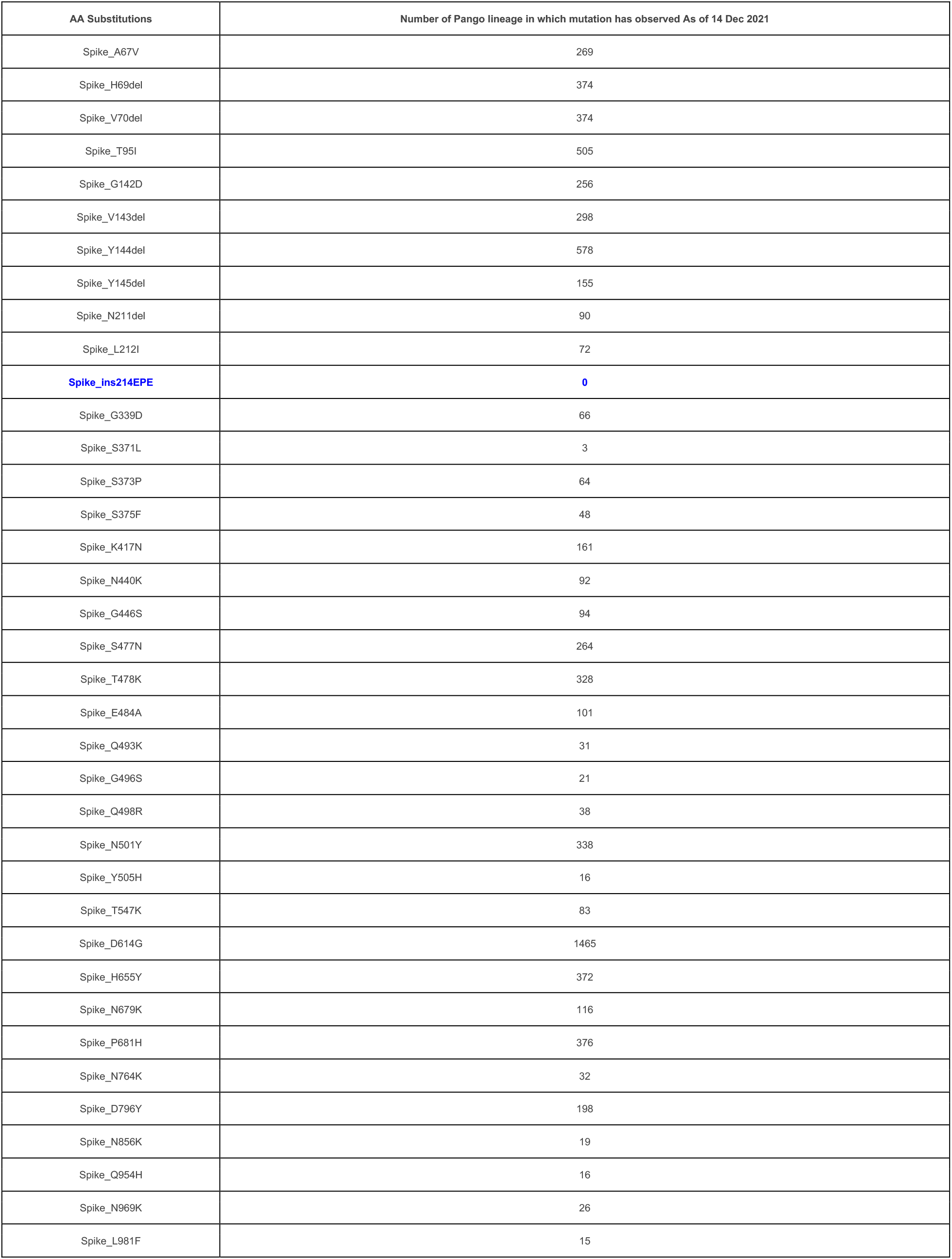
Number of PANGO lineages with mutations in the Omicron variant’s Spike protein. The Omicron sequences deposited on November 24, 2021 that were assigned B.1 are not being considered.

| AA Substitutions | Number of Pango lineage in which mutation has observed As of 14 Dec 2021 |
| --- | --- |
| Spike_A67V | 269 |
| Spike_H69del | 374 |
| Spike_V70del | 374 |
| Spike_T95I | 505 |
| Spike_G142D | 256 |
| Spike_V143del | 298 |
| Spike_Y144del | 578 |
| Spike_Y145del | 155 |
| Spike_N211del | 90 |
| Spike_L212I | 72 |
| **Spike_ins214EPE** | **0** |
| Spike_G339D | 66 |
| Spike_S371L | 3 |
| Spike_S373P | 64 |
| Spike_S375F | 48 |
| Spike_K417N | 161 |
| Spike_N440K | 92 |
| Spike_G446S | 94 |
| Spike_S477N | 264 |
| Spike_T478K | 328 |
| Spike_E484A | 101 |
| Spike_Q493K | 31 |
| Spike_G496S | 21 |
| Spike_Q498R | 38 |
| Spike_N501Y | 338 |
| Spike_Y505H | 16 |
| Spike_T547K | 83 |
| Spike_D614G | 1465 |
| Spike_H655Y | 372 |
| Spike_N679K | 116 |
| Spike_P681H | 376 |
| Spike_N764K | 32 |
| Spike_D796Y | 198 |
| Spike_N856K | 19 |
| Spike_Q954H | 16 |
| Spike_N969K | 26 |
| Spike_L981F | 15 |

**Table S2.**
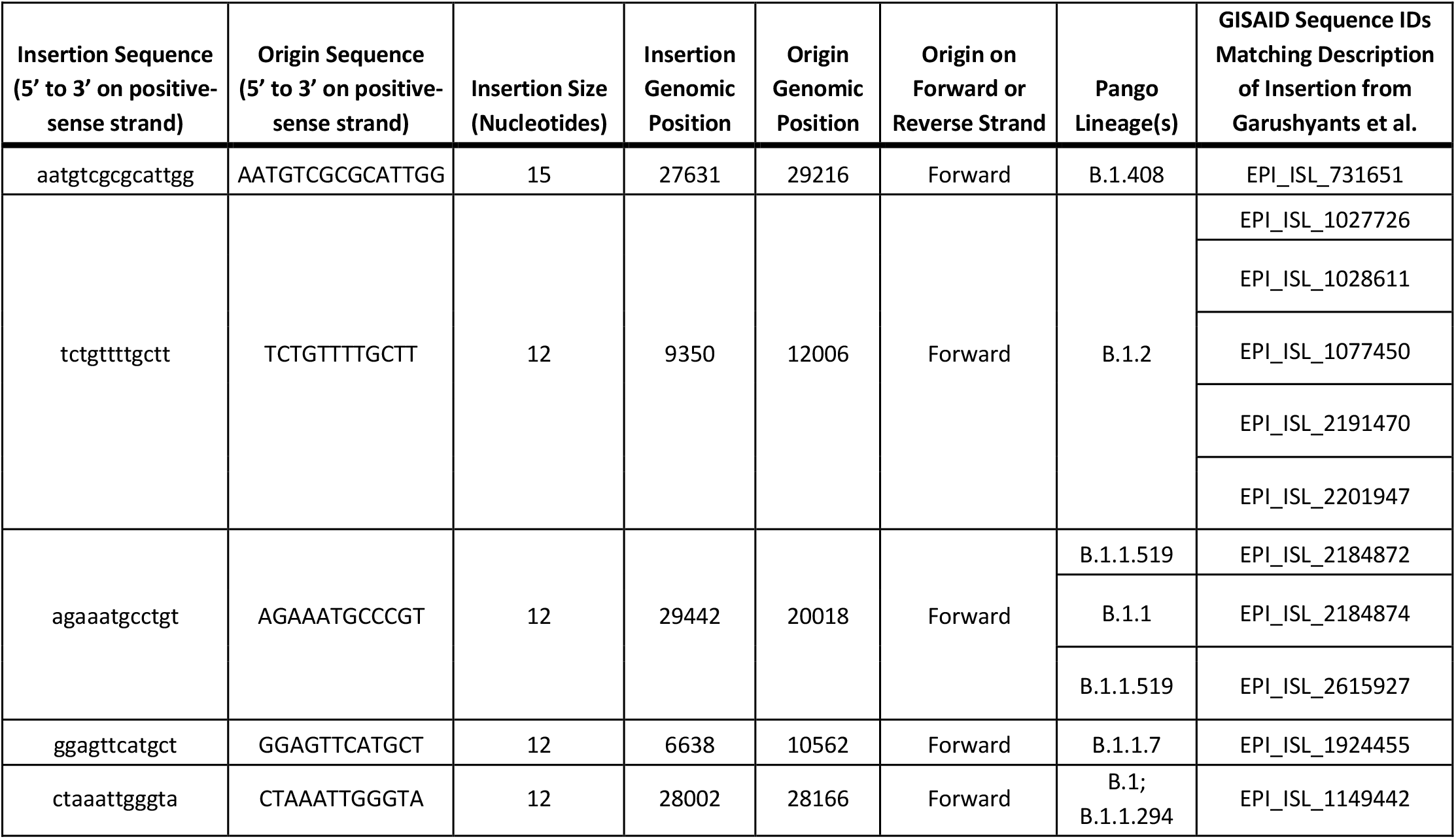
SARS-CoV-2 insertions identified and attributed to template switching in the previous analysis by Garushyants, et al. All columns except for the last column are a subset of Supplementary Data 4 from the prior analysis.^14^ With this information, we searched GISAID to identify the specific SARS-CoV-2 genomes in which the insertion and origin sequences were present at the specified locations. These genomes are identified in the last column. These events were considered as “positive controls” to evaluate whether homology in the preceding and/or subsequent nucleotide sequences are required for the generation of template switch-mediated insertions in SARS-CoV-2.

**Table S3.**
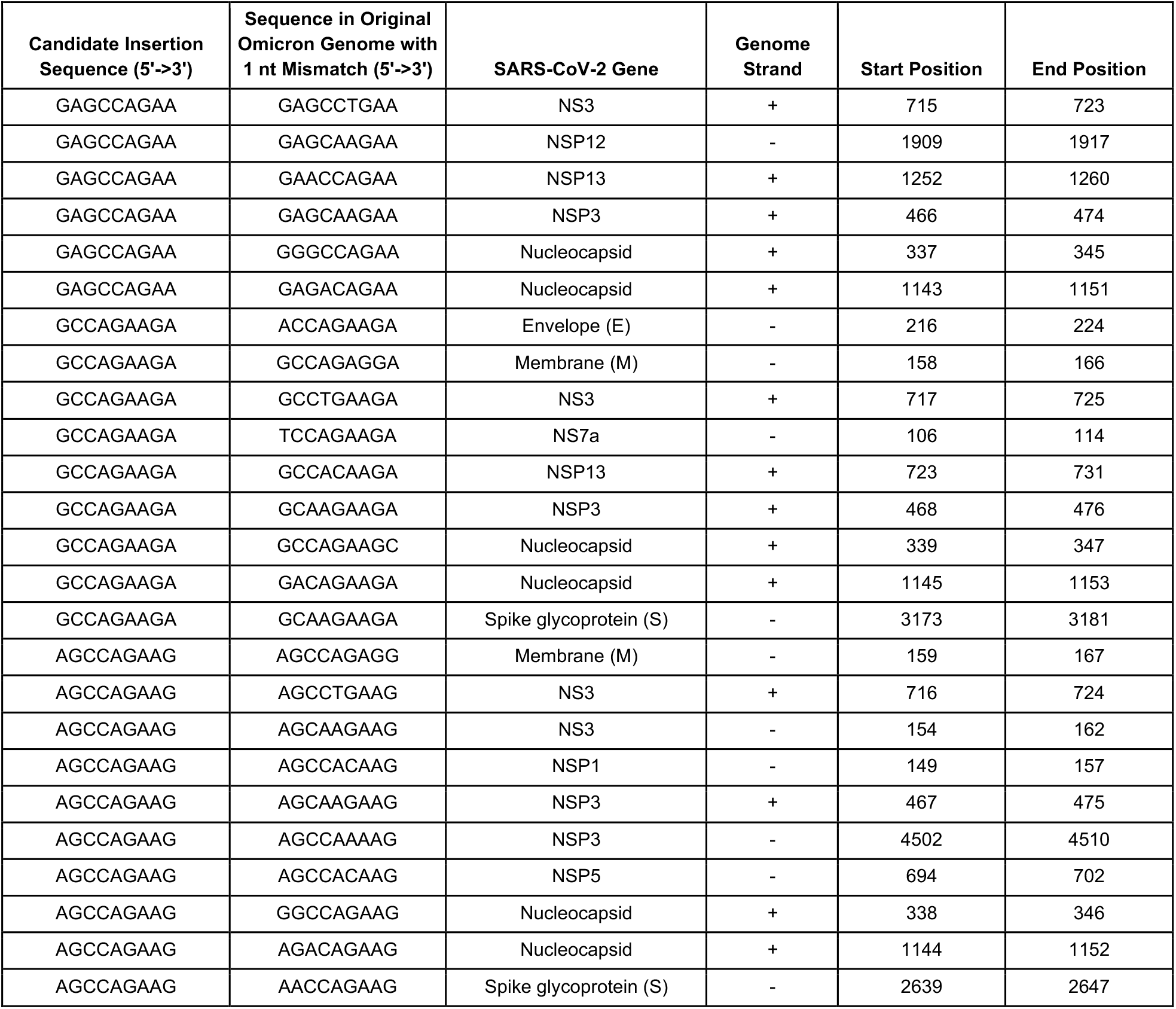
Sequences in the Omicron genome with a single nucleotide mismatch compared to the three candidate insertion sequences. The reference Omicron genome (EPI_ISL_6640916)^39^ was obtained from GISAID. Within this genome, we identified all 9-nucleotide sequences that had only one mismatch compared to the three candidate insertion sequences (5’-GAGCCAGAA-3’; 5’-AGCCAGAAG-3’; 5’-GCCAGAAGA-3’). Each hit is shown along with the gene in which it occurs, the genome strand on which it occurs (positive sense genome or negative sense anti-genome), and the within-gene start and end genomic positions. All genomic positions are shown with respect to the positive sense genome.

| Candidate Insertion Sequence (5'→3') | Sequence in Original Omicron Genome with 1 nt Mismatch (5'→3') | SARS-CoV-2 Gene | Genome Strand | Start Position | End Position |
| --- | --- | --- | --- | --- | --- |
| GAGCCAGAA | GAGCCTGAA | NS3 | + | 715 | 723 |
| GAGCCAGAA | GAGCAAGAA | NSP12 | - | 1909 | 1917 |
| GAGCCAGAA | GAACCAGAA | NSP13 | + | 1252 | 1260 |
| GAGCCAGAA | GAGCAAGAA | NSP3 | + | 466 | 474 |
| GAGCCAGAA | GGGCCAGAA | Nucleocapsid | + | 337 | 345 |
| GAGCCAGAA | GAGACAGAA | Nucleocapsid | + | 1143 | 1151 |
| GCCAGAAGA | ACCAGAAGA | Envelope (E) | - | 216 | 224 |
| GCCAGAAGA | GCCAGAGGA | Membrane (M) | - | 158 | 166 |
| GCCAGAAGA | GCCTGAAGA | NS3 | + | 717 | 725 |
| GCCAGAAGA | TCCAGAAGA | NS7a | - | 106 | 114 |
| GCCAGAAGA | GCCACAAGA | NSP13 | + | 723 | 731 |
| GCCAGAAGA | GCAAGAAGA | NSP3 | + | 468 | 476 |
| GCCAGAAGA | GCCAGAAGC | Nucleocapsid | + | 339 | 347 |
| GCCAGAAGA | GACAGAAGA | Nucleocapsid | + | 1145 | 1153 |
| GCCAGAAGA | GCAAGAAGA | Spike glycoprotein (S) | - | 3173 | 3181 |
| AGCCAGAAG | AGCCAGAGG | Membrane (M) | - | 159 | 167 |
| AGCCAGAAG | AGCCTGAAG | NS3 | + | 716 | 724 |
| AGCCAGAAG | AGCAAGAAG | NS3 | - | 154 | 162 |
| AGCCAGAAG | AGCCACAAG | NSP1 | - | 149 | 157 |
| AGCCAGAAG | AGCAAGAAG | NSP3 | + | 467 | 475 |
| AGCCAGAAG | AGCCAAAAG | NSP3 | - | 4502 | 4510 |
| AGCCAGAAG | AGCCACAAG | NSP5 | - | 694 | 702 |
| AGCCAGAAG | GGCCAGAAG | Nucleocapsid | + | 338 | 346 |
| AGCCAGAAG | AGACAGAAG | Nucleocapsid | + | 1144 | 1152 |
| AGCCAGAAG | AACCAGAAG | Spike glycoprotein (S) | - | 2639 | 2647 |

**Table S4.** Number of genomes with exact matches to candidate insertions by Pango lineage. For all three possible 9-nucleotide sequences corresponding to ins214EPE (5’-GAGCCAGAA-3’; 5’-AGCCAGAAG-3’; 5’-GCCAGAAGA-3’), we searched for SARS-CoV-2 genomes in GISAID with exact matches in the genome (positive sense) or anti-genome (negative sense). The number of such matches is shown for each Pango lineage with at least one match. Because Omicron corresponds to the Pango lineage B.1.1.529, other descendants of B.1.1 are shown in red, and B.1.1 itself is shown in red, italics, and bold.

| 5'-GAGCCAGAA-3' |  |  |  | 5'-AGCCAGAAG-3' |  |  |  | 5'-GCCAGAAGA-3' |  |  |  |
| --- | --- | --- | --- | --- | --- | --- | --- | --- | --- | --- | --- |
| Positive (+) sense | No. of genomes | Negative (-) sense | No. of genomes | Positive (+) sense | No. of genomes | Negative (-) sense | No. of genomes | Positive (+) sense | No. of genomes | Negative (-) sense | No. of genomes |
| BA.1 | 1169 | AY.44 | 13 | BA.1 | 1169 | **B.1.1.7** | **33** | BA.1 | 1169 | AY.25 | 196771 |
| AY.47 | 227 | AY.25 | 6 | B.1.177.60 | 27 | **B.1.1** | **27** | **B.1.1.7** | **28** | Unassigned | 1523 |
| AY.4.2.1 | 117 | B.1.177 | 2 | **B.1.1.7** | **16** | AY.103 | 20 | B.1.177.60 | 27 | AY.39 | 352 |
| **B.1.1.176** | **81** | **B.1.1.216** | **1** | AY.45 | 13 | AY.122 | 16 | AY.4 | 15 | AY.113 | 296 |
| AY.122 | 48 | B.1.582 | 1 | B.1.617.2 | 8 | B.1 | 13 | **B.1.1.397** | **13** | B.1.617.2 | 269 |
| AY.4 | 39 | AY.13 | 1 | AY.4 | 6 | AY.4 | 11 | AY.45 | 13 | AY.122 | 258 |
| **B.1.1.7** | **35** | AY.4 | 1 | AY.103 | 5 | AY.20 | 10 | B.1.617.2 | 9 | AY.4 | 215 |
| P.1.15 | 30 | AY.122 | 1 | Unassigned | 4 | P.1 | 10 | Unassigned | 6 | AY.25.1 | 192 |
| AY.29 | 30 | B.1.2 | 1 | P.1 | 4 | B.1.427 | 9 | AY.44 | 4 | AY.43 | 166 |
| B.1.177.60 | 27 |  |  | AY.125 | 3 | B.1.2 | 9 | P.1 | 4 | AY.44 | 147 |
| B.1.2 | 24 |  |  | AY.43 | 3 | AY.25 | 9 | AY.125 | 3 | AY.75 | 110 |
| B.1 | 23 |  |  | AY.44 | 2 | AY.39 | 7 | AY.43 | 3 | AY.46.6 | 100 |
| **B.1.1.519** | **18** |  |  | B.1.177.21 | 1 | AY.3.1 | 7 | AY.3 | 2 | **B.1.1.7** | **87** |
| AY.16 | 18 |  |  | **B.1.1.39** | **1** | AY.44 | 6 | AY.36 | 2 | AY.120 | 85 |
| AY.45 | 17 |  |  | B.1.320 | 1 | AY.43 | 6 | AY.4.2 | 2 | AY.103 | 78 |
| B.1.617.2 | 16 |  |  | B.40 | 1 | AY.3 | 5 | AY.46.5 | 1 | B | 66 |
| **B.1.1** | **14** |  |  | B.1.36.8 | 1 | AY.26 | 5 | AY.118 | 1 | AY.42 | 65 |
| AY.103 | 13 |  |  | AY.4.1 | 1 | A.21 | 5 | AY.39.1 | 1 | AY.114 | 63 |
| AY.43 | 12 |  |  | AY.46.5 | 1 | Unassigned | 4 | AY.4.2.1 | 1 | AY.33 | 55 |
| AY.46 | 11 |  |  | B | 1 | B.1.617.2 | 4 | B.1 | 1 | AY.121 | 47 |
| AY.9 | 10 |  |  | AY.4.2 | 1 | AY.9.2 | 3 | AY.25 | 1 | AY.92 | 43 |
| Unassigned | 8 |  |  | AY.36 | 1 | **B.1.1.519** | **3** | AY.33 | 1 | AY.126 | 41 |
| P.1 | 7 |  |  | **B.1.1** | **1** | B.1.258.17 | 3 | AY.122 | 1 | AY.98.1 | 38 |
| AY.44 | 7 |  |  | A.2.5 | 1 | AY.45 | 2 | AY.126 | 1 | AY.45 | 33 |
| AY.14 | 5 |  |  | B.1 | 1 | B.1.177.21 | 2 | AY.29 | 1 | AY.3 | 29 |
| AY.25 | 5 |  |  | AY.4.2.1 | 1 | AY.114 | 2 | AY.4.1 | 1 | B.1 | 27 |
| AY.113 | 4 |  |  | AY.39.1 | 1 | AY.125 | 2 | A.2.5 | 1 | AY.5.4 | 25 |
| AY.75 | 3 |  |  |  |  | B.1.609 | 2 | **B.1.1** | **1** | AY.101 | 24 |
| AY.91 | 3 |  |  |  |  | AY.61 | 2 | B.1.177.77 | 1 | AY.20 | 19 |
| AY.88 | 3 |  |  |  |  | AY.27 | 2 | B.1.36.8 | 1 | AY.88 | 18 |
| AY.127 | 3 |  |  |  |  | P.1.2 | 2 | B.1.320 | 1 | AY.99 | 18 |
| B.1.177.21 | 3 |  |  |  |  | **B.1.1.306** | **2** | **B.1.1.39** | **1** | AY.99.2 | 17 |
| AY.125 | 3 |  |  |  |  | **B.1.1.214** | **2** | B.40 | 1 | AY.108 | 15 |
| AY.3 | 2 |  |  |  |  | B.1.621 | 1 | AY.54 | 1 | AY.102 | 14 |
| AY.100 | 2 |  |  |  |  | AY.77 | 1 |  |  | AY.22 | 13 |
| B.1.429 | 2 |  |  |  |  | AY.29.1 | 1 |  |  | AY.46.4 | 12 |
| AY.121 | 2 |  |  |  |  | AY.36 | 1 |  |  | AY.3.1 | 12 |
| AY.4.2 | 2 |  |  |  |  | B.1.617.1 | 1 |  |  | AY.124 | 12 |
| B.40 | 2 |  |  |  |  | B.1.36 | 1 |  |  | B.1.2 | 11 |
| P.1.14 | 2 |  |  |  |  | AY.121 | 1 |  |  | AY.100 | 10 |
| B.1.36.39 | 2 |  |  |  |  | B.1.582 | 1 |  |  | AY.106 | 10 |
| AY.4.6 | 2 |  |  |  |  | AY.124 | 1 |  |  | C.37 | 10 |
| **B.1.1.28** | **2** |  |  |  |  | AY.98 | 1 |  |  | AY.26 | 9 |
| B.1.621.1 | 2 |  |  |  |  | **B.1.1.161** | **1** |  |  | AY.47 | 9 |
| C.30 | 2 |  |  |  |  | B.1.177 | 1 |  |  | AY.43.3 | 9 |
| AY.4.2.2 | 2 |  |  |  |  | B.1.351 | 1 |  |  | AY.82 | 7 |
| AY.46.6 | 2 |  |  |  |  | B.1.221.4 | 1 |  |  | B.1.1 | 7 |
| B.1.221 | 2 |  |  |  |  | **B.1.1.269** | **1** |  |  | AY.93 | 7 |
| AY.98 | 1 |  |  |  |  | B.1.526 | 1 |  |  | B.1.429 | 7 |
| AY.46.5 | 1 |  |  |  |  | AY.13 | 1 |  |  | AY.9.2 | 7 |
| AY.36 | 1 |  |  |  |  | **B.1.1.271** | **1** |  |  | AY.85 | 6 |
| AY.39 | 1 |  |  |  |  | **B.1.1.449** | **1** |  |  | AY.43.1 | 6 |
| AY.13 | 1 |  |  |  |  | B.1.596 | 1 |  |  | AY.119 | 5 |
| AY.122.1 | 1 |  |  |  |  | B.1.637 | 1 |  |  | AY.70 | 5 |
| B.1.177.81 | 1 |  |  |  |  | B.1.320 | 1 |  |  | AY.95 | 5 |
| B.1.351 | 1 |  |  |  |  | B.40 | 1 |  |  | B.1.582 | 5 |
| AY.39.1 | 1 |  |  |  |  | AY.39.1.1 | 1 |  |  | AY.96 | 4 |
| AY.126 | 1 |  |  |  |  |  |  |  |  | AY.5.3 | 4 |
| AY.4.1 | 1 |  |  |  |  |  |  |  |  | AY.34.1 | 4 |
| AY.5.2 | 1 |  |  |  |  |  |  |  |  | AY.4.5 | 4 |
| AY.57 | 1 |  |  |  |  |  |  |  |  | B.1.637 | 4 |
| AY.124 | 1 |  |  |  |  |  |  |  |  | AY.39.1 | 4 |
| B.1.36.8 | 1 |  |  |  |  |  |  |  |  | AY.120.1 | 3 |
| B.1.320 | 1 |  |  |  |  |  |  |  |  | AY.46 | 3 |
| B.1.1.39 | 1 |  |  |  |  |  |  |  |  | AY.1 | 3 |
| B.1.426 | 1 |  |  |  |  |  |  |  |  | AY.73 | 3 |
| B.1.147 | 1 |  |  |  |  |  |  |  |  | AY.57 | 3 |
| B.1.258 | 1 |  |  |  |  |  |  |  |  | AY.109 | 3 |
| AY.61 | 1 |  |  |  |  |  |  |  |  | AY.30 | 3 |
| B.1.575 | 1 |  |  |  |  |  |  |  |  | B.1.177 | 2 |
| B.1.1.89 | 1 |  |  |  |  |  |  |  |  | B.1.1.214 | 2 |
| AY.108 | 1 |  |  |  |  |  |  |  |  | AY.84 | 2 |
| AY.19 | 1 |  |  |  |  |  |  |  |  | AY.6 | 2 |
| AY.24 | 1 |  |  |  |  |  |  |  |  | AY.123 | 2 |
| A.2.5 | 1 |  |  |  |  |  |  |  |  | AY.77 | 2 |
| B.1.398 | 1 |  |  |  |  |  |  |  |  | AY.43.4 | 2 |
| AY.84 | 1 |  |  |  |  |  |  |  |  | AY.23 | 2 |
| B.1.561 | 1 |  |  |  |  |  |  |  |  | B.1.565 | 2 |
| AY.37 | 1 |  |  |  |  |  |  |  |  | AY.118 | 2 |
| B.1.36 | 1 |  |  |  |  |  |  |  |  | AY.125 | 2 |
| AY.98.1 | 1 |  |  |  |  |  |  |  |  | AY.19 | 2 |
| C.37 | 1 |  |  |  |  |  |  |  |  | AY.4.2.3 | 2 |
| B | 1 |  |  |  |  |  |  |  |  | AY.5 | 2 |
| AY.20 | 1 |  |  |  |  |  |  |  |  | AY.110 | 2 |
| AY.34 | 1 |  |  |  |  |  |  |  |  | AY.78 | 2 |
|  |  |  |  |  |  |  |  |  |  | B.1.1.526 | 2 |
|  |  |  |  |  |  |  |  |  |  | P.1 | 2 |
|  |  |  |  |  |  |  |  |  |  | B.1.177.21 | 2 |
|  |  |  |  |  |  |  |  |  |  | B.1.160.22 | 2 |
|  |  |  |  |  |  |  |  |  |  | AY.79 | 2 |
|  |  |  |  |  |  |  |  |  |  | B.1.596 | 2 |
|  |  |  |  |  |  |  |  |  |  | AY.54 | 2 |
|  |  |  |  |  |  |  |  |  |  | AY.111 | 2 |
|  |  |  |  |  |  |  |  |  |  | AY.59 | 2 |
|  |  |  |  |  |  |  |  |  |  | AY.61 | 1 |
|  |  |  |  |  |  |  |  |  |  | B.55 | 1 |
|  |  |  |  |  |  |  |  |  |  | AY.34.2 | 1 |
|  |  |  |  |  |  |  |  |  |  | AY.49 | 1 |
|  |  |  |  |  |  |  |  |  |  | B.1.415.1 | 1 |
|  |  |  |  |  |  |  |  |  |  | B.1.619 | 1 |
|  |  |  |  |  |  |  |  |  |  | AY.24 | 1 |
|  |  |  |  |  |  |  |  |  |  | AY.14 | 1 |
|  |  |  |  |  |  |  |  |  |  | B.1.254 | 1 |
|  |  |  |  |  |  |  |  |  |  | AY.53 | 1 |
|  |  |  |  |  |  |  |  |  |  | B.1.581 | 1 |
|  |  |  |  |  |  |  |  |  |  | B.1.459 | 1 |
|  |  |  |  |  |  |  |  |  |  | AY.98 | 1 |
|  |  |  |  |  |  |  |  |  |  | AY.4.2 | 1 |
|  |  |  |  |  |  |  |  |  |  | AY.67 | 1 |
|  |  |  |  |  |  |  |  |  |  | AY.4.2.1 | 1 |
|  |  |  |  |  |  |  |  |  |  | AY.18 | 1 |
|  |  |  |  |  |  |  |  |  |  | AY.32 | 1 |
|  |  |  |  |  |  |  |  |  |  | AY.116 | 1 |
|  |  |  |  |  |  |  |  |  |  | B.1.320 | 1 |
|  |  |  |  |  |  |  |  |  |  | AY.117 | 1 |
|  |  |  |  |  |  |  |  |  |  | AY.7.2 | 1 |
|  |  |  |  |  |  |  |  |  |  | AY.13 | 1 |
|  |  |  |  |  |  |  |  |  |  | C.37.1 | 1 |
|  |  |  |  |  |  |  |  |  |  | AY.58 | 1 |
|  |  |  |  |  |  |  |  |  |  | AY.127 | 1 |
|  |  |  |  |  |  |  |  |  |  | B.1.466.2 | 1 |
|  |  |  |  |  |  |  |  |  |  | AY.90 | 1 |
|  |  |  |  |  |  |  |  |  |  | B.1.535 | 1 |
|  |  |  |  |  |  |  |  |  |  | AY.76 | 1 |
|  |  |  |  |  |  |  |  |  |  | P.2 | 1 |
|  |  |  |  |  |  |  |  |  |  | AY.46.2 | 1 |
|  |  |  |  |  |  |  |  |  |  | B.1.1.519 | 1 |
|  |  |  |  |  |  |  |  |  |  | A.2.5 | 1 |
|  |  |  |  |  |  |  |  |  |  | B.1.177.81 | 1 |
|  |  |  |  |  |  |  |  |  |  | B.1.36.38 | 1 |
|  |  |  |  |  |  |  |  |  |  | P.1.1 | 1 |
|  |  |  |  |  |  |  |  |  |  | B.1.469 | 1 |
|  |  |  |  |  |  |  |  |  |  | AY.27 | 1 |
|  |  |  |  |  |  |  |  |  |  | B.1.1.449 | 1 |
|  |  |  |  |  |  |  |  |  |  | B.1.1.153 | 1 |
|  |  |  |  |  |  |  |  |  |  | AY.69 | 1 |
|  |  |  |  |  |  |  |  |  |  | AY.50 | 1 |
|  |  |  |  |  |  |  |  |  |  | AY.29 | 1 |
|  |  |  |  |  |  |  |  |  |  | B.1.1.271 | 1 |
|  |  |  |  |  |  |  |  |  |  | B.1.621 | 1 |
|  |  |  |  |  |  |  |  |  |  | AY.39.1.1 | 1 |

**Table S5.** Number of seasonal or enteric *Coronaviridae* genomes with exact matches to each candidate insertion sequence. We searched non-SARS-CoV-2 *Coronaviridae* genomes and anti-genomes for exact matches to all three possible 9-nucleotide sequences (5’-GAGCCAGAA-3’; 5’-AGCCAGAAG-3’; 5’-GCCAGAAGA-3’) that gave rise to ins214EPE in Omicron. This table shows the number of matches that were identified for each such virus-gene combination with at least one match, along with the strand (positive sense genome or negative sense anti-genome) on which the match was found.

| Candidate Insertion Sequence (5'->3') | Genome Strand | Virus | Gene | Number of Genomes with Match |
| --- | --- | --- | --- | --- |
| GCCAGAAGA | + | Human coronavirus OC43 | Nucleocapsid (N) | 177 |
| GCCAGAAGA | + | Human enteric coronavirus strain 4408 | Nucleocapsid (N) | 2 |
| GCCAGAAGA | + | Human coronavirus OC43 | Protein I (internal to N) | 6 |
| GAGCCAGAA | + | Human coronavirus OC43 | Replicase polyprotein 1a (ORF1a) | 98 |
| GAGCCAGAA | + | Human enteric coronavirus strain 4408 | Replicase polyprotein 1a (ORF1a) | 1 |
| GAGCCAGAA | - | Human coronavirus 229E | Spike glycoprotein (S) | 33 |
| GCCAGAAGA | - | Human coronavirus NL63 | Spike glycoprotein (S) | 2 |

**Table S6.**
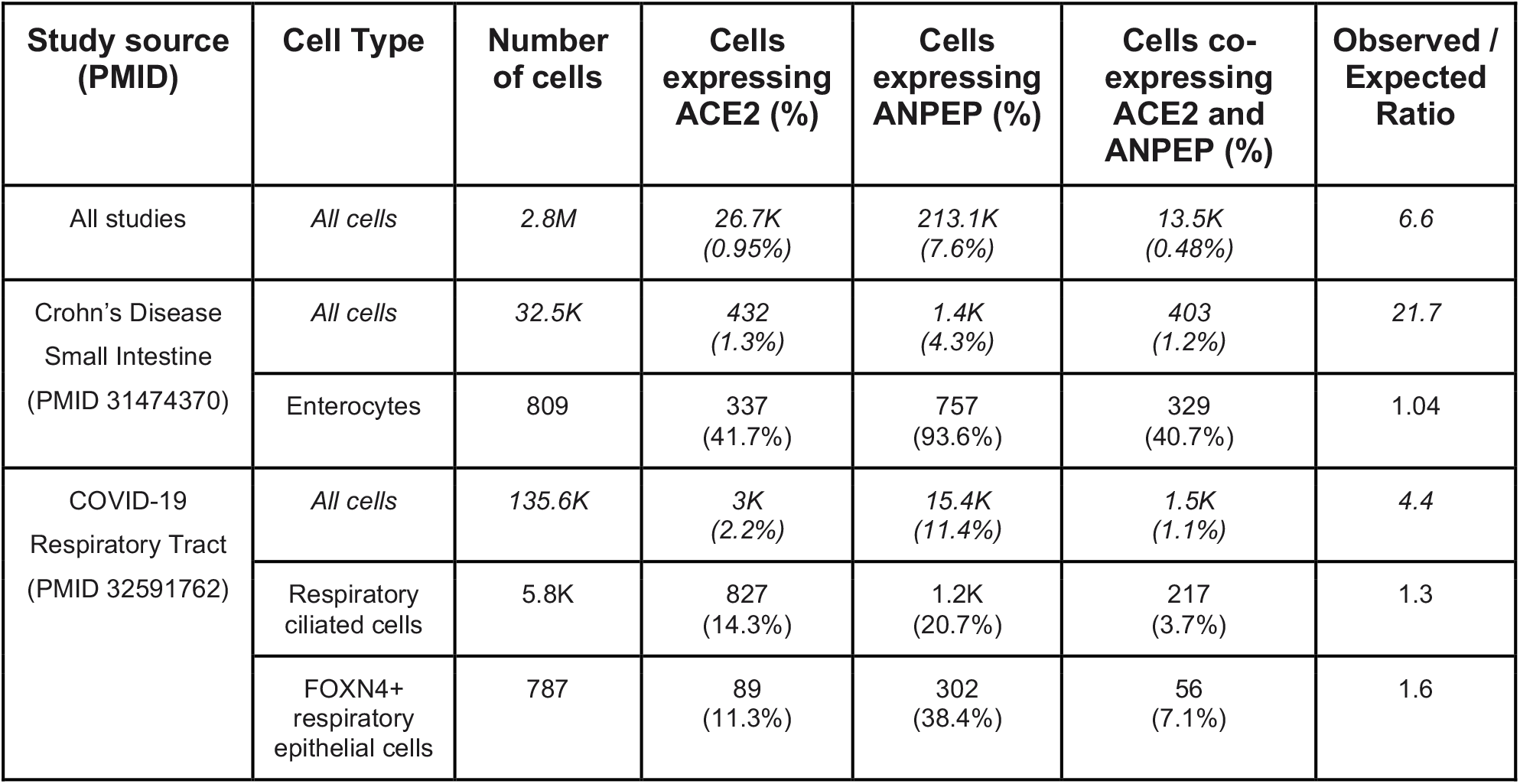
Coexpression analysis of ACE2 and ANPEP in single cell RNA-sequencing datasets. The number and percent of cells in each category expressing ACE2, ANPEP, or both ACE2 and ANPEP are shown, along with the observed to expected ratio of co-expression for the given category. In the first row, “All studies” corresponds to the set of studies that are hosted in the previously described Single Cell application at academia.nferx.com.^46,76^ The analyzed enterocytes are derived from a study of ileal biopsies from Crohn’s Disease patients.^78^ The analyzed respiratory ciliated cells and FOXN4+ respiratory epithelial cells are derived from a study of nasopharyngeal and bronchial samples from COVID-19 patients and healthy controls.^77^

**Table S7.** Coexpression analysis of ACE2 and DPP4 in single cell RNA-sequencing datasets. The number and percent of cells in each category expressing ACE2, DPP4, or both ACE2 and DPP4 are shown, along with the observed to expected ratio of co-expression for the given category. In the first row, “All studies” corresponds to the set of studies that are hosted in the previously described Single Cell application at academia.nferx.com.^46,76^ The analyzed enterocytes are derived from a study of ileal biopsies from Crohn’s Disease patients.^78^ The analyzed respiratory ciliated cells and FOXN4+ respiratory epithelial cells are derived from a study of nasopharyngeal and bronchial samples from COVID-19 patients and healthy controls.^77^

| Study source (PMID) | Cell Type | Number of cells | Cells expressing ACE2 (%) | Cells expressing DPP4 (%) | Cells co-expressing ACE2 and DPP4 (%) | Observed / Expected Ratio |
| --- | --- | --- | --- | --- | --- | --- |
| All studies | All cells | 2.8M | 26.7K<br>(0.95%) | 112.5K<br>(4.02%) | 6.4K<br>(0.23%) | 6.0 |
| Crohn's Disease<br>Small Intestine<br>(PMID 31474370) | All cells | 32.5K | 432<br>(1.3%) | 1.3K<br>(4.9%) | 175<br>(0.54%) | 10.1 |
|  | Enterocytes | 809 | 337<br>(41.7%) | 254<br>(31.4%) | 153<br>(18.9%) | 1.4 |

**Table S8.**
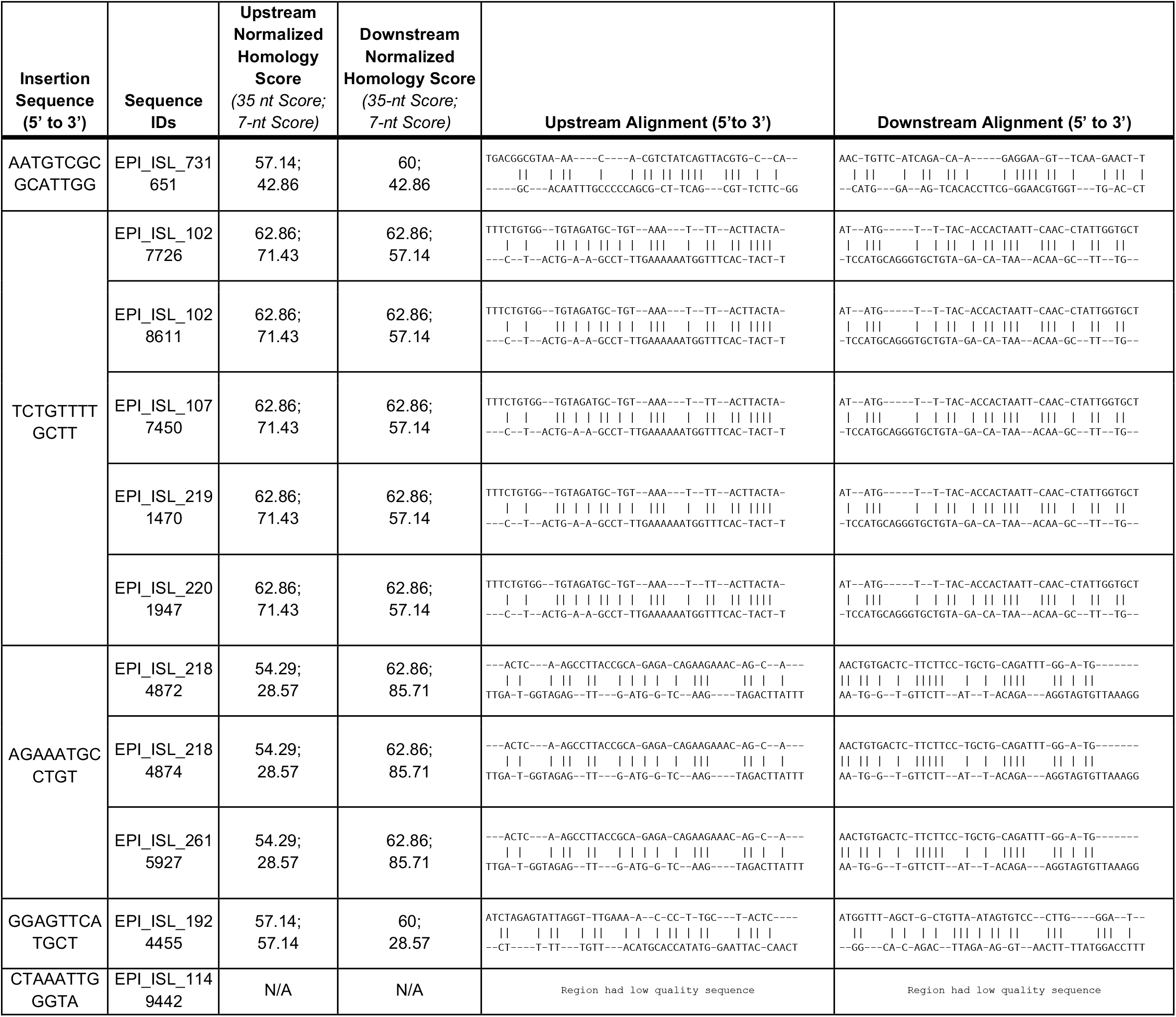
Local alignment and homology scores for nucleotides upstream or downstream of origin and insertion sites from previously identified template switch-mediated insertions. These correspond to five “positive control” SARS-CoV-2 template switch-mediated insertions shown in Table S6, which were identified in the prior analysis by Garushyants, et al.^14^ We identified the SARS-CoV-2 genomes in GISAID corresponding to each insertion, and then calculated normalized homology scores (NHS) for the 35 or 7 nucleotides upstream or downstream from each insertion and origin sequence. The scores along with the alignments are shown here, and the scores are compared to a null distribution of NHS values computed between 10,000 pairs of randomly selected non-overlapping 35- or 7-nucleotide sequences from the reference SARS-CoV-2 genome in **Figure S3**.

## References

1. Classification of Omicron (B.1.1.529): SARS-CoV-2 Variant of Concern. https://www.who.int/news/item/26-11-2021-classification-of-omicron-(b.1.1.529)-sars-cov-2-variant-of-concern.

2. Shu, Y. & McCauley, J. GISAID: Global initiative on sharing all influenza data - from vision to reality. Euro Surveill. 22, (2017).

3. GISAID - hCov19 Variants. https://www.gisaid.org/hcov19-variants/.

4. Plante, J. A. et al. Spike mutation D614G alters SARS-CoV-2 fitness. Nature 592, 116–121 (2020).

5. Daniloski, Z. et al. The Spike D614G mutation increases SARS-CoV-2 infection of multiple human cell types. (2021) doi:10.7554/eLife.65365.

6. Wang, P. et al. Antibody resistance of SARS-CoV-2 variants B.1.351 and B.1.1.7. Nature (2021) doi:10.1038/s41586-021-03398-2.

7. Uriu, K. et al. Neutralization of the SARS-CoV-2 Mu Variant by Convalescent and Vaccine Serum. N. Engl. J. Med. (2021) doi:10.1056/NEJMc2114706.

8. Collier, D. A. et al. Sensitivity of SARS-CoV-2 B.1.1.7 to mRNA vaccine-elicited antibodies. Nature 593, 136–141 (2021).

9. McCarthy, K. R. et al. Recurrent deletions in the SARS-CoV-2 spike glycoprotein drive antibody escape. Science 371, 1139–1142 (2021).

10. Motozono, C. et al. SARS-CoV-2 spike L452R variant evades cellular immunity and increases infectivity. Cell Host Microbe 29, (2021).

11. McCallum, M. et al. N-terminal domain antigenic mapping reveals a site of vulnerability for SARS-CoV-2. Cell 184, 2332–2347.e16 (2021).

12. Harvey, W. T. et al. SARS-CoV-2 variants, spike mutations and immune escape. Nat. Rev. Microbiol. 19, 409–424 (2021).

13. Ku, Z. et al. Molecular determinants and mechanism for antibody cocktail preventing SARS-CoV-2 escape. Nat. Commun. 12, 1–13 (2021).

14. Garushyants, S. K., Rogozin, I. B. & Koonin, E. V. Template switching and duplications in SARS-CoV-2 genomes give rise to insertion variants that merit monitoring. Communications Biology 4, 1–9 (2021).

15. Anand, P., Puranik, A., Aravamudan, M., Venkatakrishnan, A. J. & Soundararajan, V. SARS-CoV-2 strategically mimics proteolytic activation of human ENaC. Elife 9, (2020).

16. Coutard, B. et al. The spike glycoprotein of the new coronavirus 2019-nCoV contains a furin-like cleavage site absent in CoV of the same clade. Antiviral Res. 176, (2020).

17. Jaimes, J. A., Millet, J. K. & Whittaker, G. R. Proteolytic Cleavage of the SARS-CoV-2 Spike Protein and the Role of the Novel S1/S2 Site. iScience 23, (2020).

18. Peacock, T. P. et al. The furin cleavage site in the SARS-CoV-2 spike protein is required for transmission in ferrets. Nature Microbiology 6, 899–909 (2021).

19. Structure, Function, and Antigenicity of the SARS-CoV-2 Spike Glycoprotein. Cell 181, 281–292.e6 (2020).

20. Liu, Z. et al. Lymphocyte subset (CD4+, CD8+) counts reflect the severity of infection and predict the clinical outcomes in patients with COVID-19. J. Infect. 81, 318 (2020).

21. Diao, B. et al. Reduction and Functional Exhaustion of T Cells in Patients With Coronavirus Disease 2019 (COVID-19). Front. Immunol. 11, (2020).

22. Tavakolpour, S., Rakhshandehroo, T., Wei, E. X. & Rashidian, M. Lymphopenia during the COVID-19 infection: What it shows and what can be learned. Immunol. Lett. 225, 31 (2020).

23. Zhang, Z. et al. SARS-CoV-2 spike protein dictates syncytium-mediated lymphocyte elimination. Cell Death Differ. 28, (2021).

24. Zhang, L. et al. Reverse-transcribed SARS-CoV-2 RNA can integrate into the genome of cultured human cells and can be expressed in patient-derived tissues. Proc. Natl. Acad. Sci. U. S. A. 118, (2021).

25. Parry, R., Gifford, R. J., Lytras, S., Ray, S. C. & Coin, L. J. M. No evidence of SARS-CoV-2 reverse transcription and integration as the origin of chimeric transcripts in patient tissues. Proceedings of the National Academy of Sciences of the United States of America vol. 118 (2021).

26. Zhang, L. et al. Response to Parry et al.: Strong evidence for genomic integration of SARS-CoV-2 sequences and expression in patient tissues. Proceedings of the National Academy of Sciences of the United States of America vol. 118 (2021).

27. Tzou, P. L. et al. Coronavirus Antiviral Research Database (CoV-RDB): An Online Database Designed to Facilitate Comparisons between Candidate Anti-Coronavirus Compounds. Viruses 12, (2020).

28. Venkatakrishnan, A. J. et al. Antigenic minimalism of SARS-CoV-2 is linked to surges in COVID-19 community transmission and vaccine breakthrough infections. medRxiv 2021.05.23.21257668 (2021).

29. Gerdol, M., Dishnica, K. & Giorgetti, A. Emergence of a recurrent insertion in the N-terminal domain of the SARS-CoV-2 spike glycoprotein. bioRxiv 2021.04.17.440288 (2021) doi:10.1101/2021.04.17.440288.

30. Tarke, A. et al. Comprehensive analysis of T cell immunodominance and immunoprevalence of SARS-CoV-2 epitopes in COVID-19 cases. Cell Rep Med 2, 100204 (2021).

31. Lam, S. D., Waman, V. P., Orengo, C. & Lees, J. Insertions in the SARS-CoV-2 Spike N-Terminal Domain May Aid COVID-19 Transmission. bioRxiv 2021.12.06.471394 (2021) doi:10.1101/2021.12.06.471394.

32. Huston, N. C. et al. Comprehensive in vivo secondary structure of the SARS-CoV-2 genome reveals novel regulatory motifs and mechanisms. Mol. Cell 81, 584 (2021).

33. te Velthuis, A. J. W., Arnold, J. J., Cameron, C. E., van den Worm, S. H. E. & Snijder, E. J. The RNA polymerase activity of SARS-coronavirus nsp12 is primer dependent. Nucleic Acids Res. 38, 203–214 (2009).

34. Kim, D. et al. The Architecture of SARS-CoV-2 Transcriptome. Cell 181, (2020).

35. Jackson, B. et al. Generation and transmission of interlineage recombinants in the SARS-CoV-2 pandemic. Cell 184, 5179–5188.e8 (2021).

36. Sawicki, S. G. & Sawicki, D. L. Coronaviruses use discontinuous extension for synthesis of subgenome-length negative strands. Adv. Exp. Med. Biol. 380, (1995).

37. Simon-Loriere, E. & Holmes, E. C. Why do RNA viruses recombine? Nat. Rev. Microbiol. 9, 617–626 (2011).

38. thomasppeacock. Putative host origins of RNA insertions in SARS-CoV-2 genomes. https://virological.org/t/putative-host-origins-of-rna-insertions-in-sars-cov-2-genomes/761 (2021).

39. Kandeel, M., Mohamed, M. E. M., Hm, A. E.-L., Venugopala, K. N. & El-Beltagi, H. S. Omicron variant genome evolution and phylogenetics. J. Med. Virol. (2021) doi:10.1002/jmv.27515.

40. Hadfield, J. et al. Nextstrain: real-time tracking of pathogen evolution. Bioinformatics vol. 34 4121–4123 (2018).

41. Turkahia, Y. et al. Pandemic-Scale Phylogenomics Reveals Elevated Recombination Rates in the SARS-CoV-2 Spike Region. bioRxiv 2021.08.04.455157 (2021) doi:10.1101/2021.08.04.455157.

42. Morens, D. M., Taubenberger, J. K. & Fauci, A. S. Universal Coronavirus Vaccines - An Urgent Need. N. Engl. J. Med. (2021) doi:10.1056/NEJMp2118468.

43. Lau, S. K. P. et al. Molecular Evolution of Human Coronavirus 229E in Hong Kong and a Fatal COVID-19 Case Involving Coinfection with a Novel Human Coronavirus 229E Genogroup. mSphere 6, (2021).

44. Kim, D., Quinn, J., Pinsky, B., Shah, N. H. & Brown, I. Rates of Co-infection Between SARS-CoV-2 and Other Respiratory Pathogens. JAMA 323, 2085–2086 (2020).

45. Musuuza, J. S. et al. Prevalence and outcomes of co-infection and superinfection with SARS-CoV-2 and other pathogens: A systematic review and meta-analysis. PLoS One 16, e0251170 (2021).

46. Venkatakrishnan, A. J. et al. Knowledge synthesis of 100 million biomedical documents augments the deep expression profiling of coronavirus receptors. Elife 9, (2020).

47. Zhou, J. et al. Human intestinal tract serves as an alternative infection route for Middle East respiratory syndrome coronavirus. Science advances 3, (2017).

48. Lamers, M. M. et al. SARS-CoV-2 productively infects human gut enterocytes. Science 369, (2020).

49. Bein, A. et al. Enteric Coronavirus Infection and Treatment Modeled With an Immunocompetent Human Intestine-On-A-Chip. Front. Pharmacol. 0, (2021).

50. Sola, I., Almazán, F., Zúñiga, S. & Enjuanes, L. Continuous and Discontinuous RNA Synthesis in Coronaviruses. (2015) doi:10.1146/annurev-virology-100114-055218.

51. Yang, Y., Yan, W., Hall, A. B. & Jiang, X. Characterizing Transcriptional Regulatory Sequences in Coronaviruses and Their Role in Recombination. Mol. Biol. Evol. 38, 1241–1248 (2020).

52. Nomburg, J., Meyerson, M. & DeCaprio, J. A. Pervasive generation of non-canonical subgenomic RNAs by SARS-CoV-2. Genome Med. 12, (2020).

53. HKUMed finds Omicron SARS-CoV-2 can infect faster and better than Delta in human bronchus but with less severe infection in lung. https://www.med.hku.hk/en/news/press/20211215-omicron-sars-cov-2-infection.

54. Callaway, E. & Ledford, H. How bad is Omicron? What scientists know so far. Nature 600, 197–199 (2021).

55. Garcia-Beltran, W. F. et al. mRNA-based COVID-19 vaccine boosters induce neutralizing immunity against SARS-CoV-2 Omicron variant. medRxiv 2021.12.14.21267755 (2021).

56. Doria-Rose, N. A. et al. Booster of mRNA-1273 Vaccine Reduces SARS-CoV-2 Omicron Escape from Neutralizing Antibodies. medRxiv 2021.12.15.21267805 (2021).

57. Dejnirattisai, W. et al. Reduced neutralisation of SARS-COV-2 Omicron-B.1.1.529 variant by post-immunisation serum. medRxiv 2021.12.10.21267534 (2021).

58. Wilhelm, A. et al. Reduced Neutralization of SARS-CoV-2 Omicron Variant by Vaccine Sera and Monoclonal Antibodies. medRxiv 2021.12.07.21267432 (2021).

59. Pfizer and BioNTech Provide Update on Omicron Variant. https://www.pfizer.com/news/press-release/press-release-detail/pfizer-and-biontech-provide-update-omicron-variant.

60. Liu, L. et al. Striking Antibody Evasion Manifested by the Omicron Variant of SARS-CoV-2. bioRxiv 2021.12.14.472719 (2021) doi:10.1101/2021.12.14.472719.

61. Zhang, L. et al. The significant immune escape of pseudotyped SARS-CoV-2 Variant Omicron. Emerg. Microbes Infect. (2021) doi:10.1080/22221751.2021.2017757.

62. Pulliam, J. R. C. et al. Increased risk of SARS-CoV-2 reinfection associated with emergence of the Omicron variant in South Africa. medRxiv 2021.11.11.21266068 (2021).

63. Varrelman, T. J., Rader, B. M., Astley, C. M. & Brownstein, J. S. Syndromic Surveillance-Based Estimates of Vaccine Efficacy Against COVID-Like Illness from Emerging Omicron and COVID-19 Variants. medRxiv 2021.12.17.21267995 (2021).

64. News room. https://www.discovery.co.za/corporate/news-room.

65. Snijder, E. J. et al. A unifying structural and functional model of the coronavirus replication organelle: Tracking down RNA synthesis. PLoS Biol. 18, (2020).

66. Kupferschmidt, K. Where did ‘weird’ Omicron come from? Science 374, (2021).

67. Wei, C. et al. Evidence for a mouse origin of the SARS-CoV-2 Omicron variant. J. Genet. Genomics (2021) doi:10.1016/j.jgg.2021.12.003.

68. Gribble, J. et al. The coronavirus proofreading exoribonuclease mediates extensive viral recombination. PLoS Pathog. 17, e1009226 (2021).

69. Madhi, S. A. et al. Population Immunity and Covid-19 Severity with Omicron Variant in South Africa. N. Engl. J. Med. (2022) doi:10.1056/NEJMoa2119658.

70. Wolter, N. et al. Early assessment of the clinical severity of the SARS-CoV-2 omicron variant in South Africa: a data linkage study. Lancet 399, (2022).

71. Ulloa, A. C., Buchan, S. A., Daneman, N. & Brown, K. A. Estimates of SARS-CoV-2 Omicron Variant Severity in Ontario, Canada. JAMA (2022) doi:10.1001/jama.2022.2274.

72. Maslo, C. et al. Characteristics and Outcomes of Hospitalized Patients in South Africa During the COVID-19 Omicron Wave Compared With Previous Waves. JAMA 327, 583–584 (2022).

73. Mathieu, E. et al. A global database of COVID-19 vaccinations. Nat Hum Behav 5, 947–953 (2021).

74. Frankish, A. et al. GENCODE 2021. Nucleic Acids Res. 49, D916–D923 (2021).

75. Brister, J. R., Ako-Adjei, D., Bao, Y. & Blinkova, O. NCBI viral genomes resource. Nucleic Acids Res. 43, D571–7 (2015).

76. Doddahonnaiah, D. et al. A Literature-Derived Knowledge Graph Augments the Interpretation of Single Cell RNA-seq Datasets. Genes 12, (2021).

77. Chua, R. L. et al. COVID-19 severity correlates with airway epithelium-immune cell interactions identified by single-cell analysis. Nat. Biotechnol. 38, (2020).

78. Martin, J. C. et al. Single-Cell Analysis of Crohn’s Disease Lesions Identifies a Pathogenic Cellular Module Associated with Resistance to Anti-TNF Therapy. Cell 178, (2019).

79. Severe acute respiratory syndrome coronavirus 2 isolate Wuhan-Hu-1, co - Nucleotide - NCBI. https://www.ncbi.nlm.nih.gov/nuccore/NC_045512.

